# Safety and Efficacy of iPSC-Derived GABAergic Interneurons for Unilateral MTLE

**DOI:** 10.64898/2026.04.10.26350582

**Authors:** Boyu Tang, Jiemin Zhou

## Abstract

**Importance:** Epilepsy is one of the most common neurological disorders globally. A significant proportion of patients fail to achieve effective seizure control with medication and ultimately develop drug-resistant epilepsy, particularly mesial temporal lobe epilepsy (MTLE). While surgical resection and laser interstitial thermal therapy (LITT) are effective treatments for drug-resistant MTLE, these procedures may be associated with severe adverse events. In contrast, allogeneic induced pluripotent stem cell (iPSC)-based therapy is expected to offer a novel, potentially safer therapeutic approach with fewer side effects for patients with drug-resistant MTLE.

**Objective:** To evaluate the safety and preliminary efficacy of a single intracranial injection of ALC05 (iPSC-derived GABAergic interneurons) in patients with unilateral MTLE, and to assess the therapeutic effects of different dosage levels.

**Design, Setting, and Participants:** This single-center, randomized, double-blind, Phase 1 clinical trial will enroll 12 subjects with unilateral MTLE. All subjects will be randomly assigned to either the low-dose or high-dose group in a 1:1 ratio. To minimize risks at each dose level, the first subject in each dose group will be monitored for safety for at least 3 months following ALC05 injection and must demonstrate acceptable safety and tolerability before the remaining subjects are enrolled. The primary outcome will be the incidence and severity of adverse events (AEs) and serious adverse events (SAEs). Secondary outcomes include cell engraftment and survival, responder rate, and seizure frequency. The follow-up period for this study is 1 year. After completing the follow-up period within this study, subjects will enter a 15-year long-term safety follow-up.

**Discussion:** MTLE remains a significant challenge in neurology. The results of this study will provide critical data regarding the feasibility and preliminary efficacy of ALC05 in treating MTLE and may offer a transformative therapeutic option for this condition.

## Introduction

Epilepsy remains a globally prevalent chronic neurological disorder, with 2021 estimates placing the disease burden at approximately 51.7 million individuals worldwide.^1^ The clinical significance of this disease extends beyond its associated mortality risks, it further precipitates substantial declines in psychosocial function, economic stability and overall quality of life.

Focal epilepsy accounts for roughly 61% of total incidence, with the majority of these cases originating in the temporal lobe.^2–4^ The most frequent subtype MTLE presents a great clinical challenge, as approximately 70% of patients develop pharmacoresistance.^5^ Surgical resection or laser ablation remain effective options for drug-resistant MTLE, ^6–8^ though both carry risk of memory impairment, language dysfunction and cognitive decline.^9^ A treatment strategy that avoids these neurological trade-offs while maintaining seizure control remains unavailable.

The core pathophysiology of epilepsy involves an imbalance between neuronal excitation and inhibition. As the principal inhibitory neurotransmitter in the brain, dysfunction of gamma-aminobutyric acid (GABA) contributes to both disease pathogenesis and progression.^10,11^ Pallial GABAergic interneurons (pINs) are the primary source of inhibition in the neocortex and hippocampus, where they modulate local circuit excitability via GABA release.^12^ Abnormalities of pINs, especially those derived from the medial ganglionic eminence (MGE), have been observed in various epilepsy cases. In animal models, MGE-pINs are markedly reduced in number or functionally impaired.^13,14^ Their selective activation has been showed to attenuate seizure activity.^15,16^ Hippocampal specimens from MTLE patients show a similar pattern of MGE-pINs depletion.^17,18^

Stem cell therapy offers significant therapeutic potential for epilepsy. Transplantation of pINs into epileptic brain regions to reinforce inhibitory function has been a promising therapeutic strategy. In rodent epilepsy models, MGE-pINs derived from embryonic tissue migrated locally and survived long-term.^19^ They matured into functional interneurons and integrated into host neural circuits. In several studies, this let to significant seizure suppression.^20–23^ Conversely, caudal ganglionic eminence pINs fail to suppress seizures and may induce disinhibition.^24^ IPSCs provide robust self-replication potential and multipotent differentiation. Under specific conditions, these cells can be directed into MGE-pINs. This approach represents a novel, minimally invasive strategy to correct circuit abnormalities and restore neural regulation in MTLE.

This study primarily evaluates the safety of single-dose stereotactic intracranial injection of ALC05 in patients with unilateral drug-resistant MTLE. Secondary objectives include assessing preliminary efficacy and exploring the clinical effects of distinct dose cohorts.

## Methods

### Study Design and Setting

This is a single-center, randomized, double-blind, phase 1 clinical trial that includes 12 patients with unilateral MTLE. The study follows the Standard Protocol Items: Recommendations for Interventional Trials (SPIRIT) reporting guideline. The complete trial protocol is available in the Supplement 1.

The study is being conducted between 2026 and 2027 in the First Affiliated Hospital of Xi’an Jiaotong University, Shaanxi, China. Potential participants will be screened for eligibility. All enrolled subjects will be randomly divided into either low dose or high dose group at a 1:1 ratio. To minimize risks to subjects at each dose level, the first subject in each dose group will be monitored for safety for at least 3 months post ALC05 injection and show acceptable safety and tolerability, before the remaining patients can be enrolled (Figure 1). The safety data will be reviewed by the Independent Data Safety Monitoring Board (DSMB). The primary endpoint is the incidence of treatment-emergent adverse events (TEAEs). Secondary endpoints include imaging evidence of cell engraftment and survival, neurocognitive and psychological function changes, and absolute changes in seizure frequency from baseline.

**Fig. 1.**
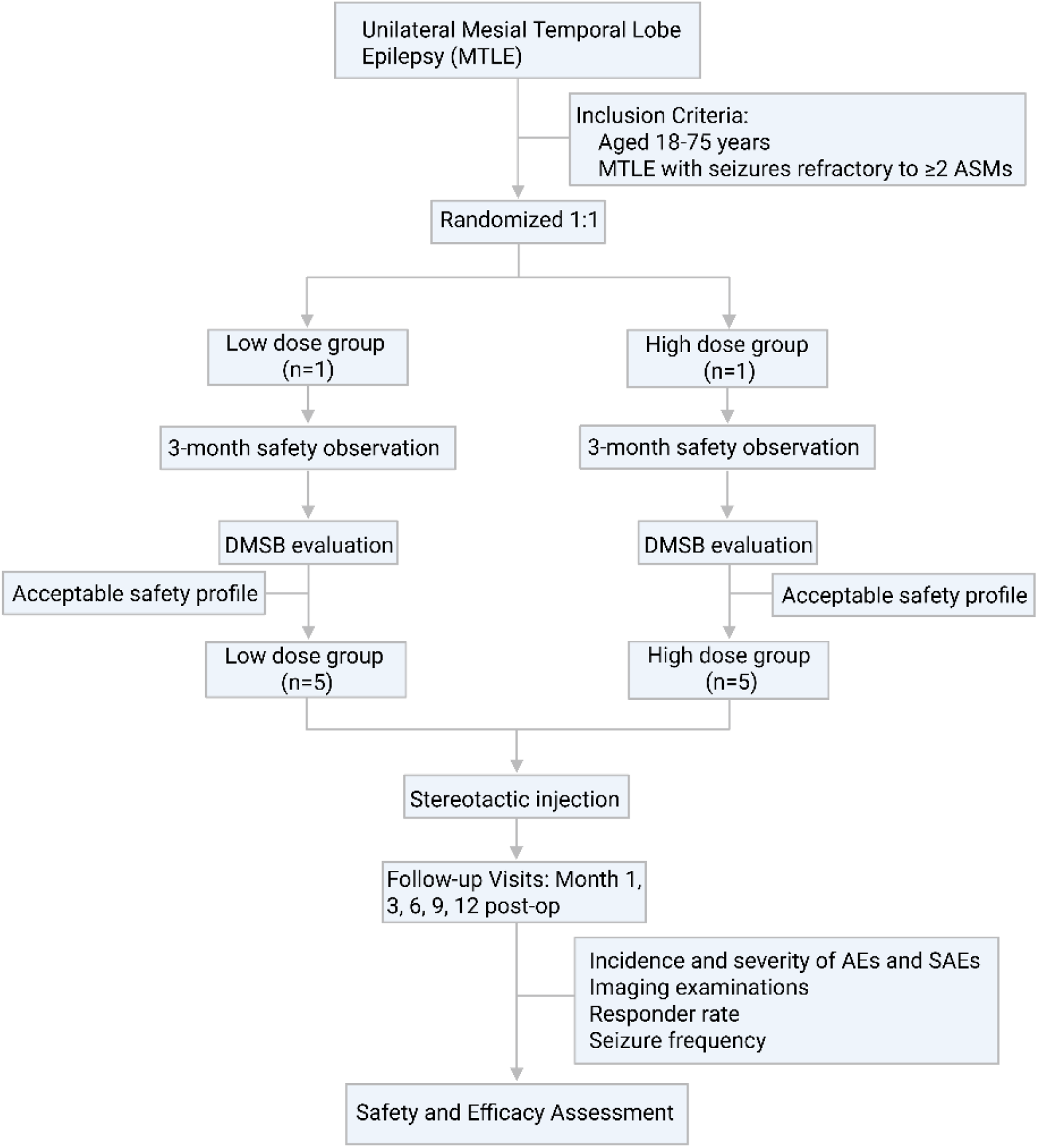
Flow Diagram.

### Eligibility Criteria

Inclusion criteria include (1) age 18 to 75 years; (2) confirmed diagnosis of drug-resistant MTLE for at least 5 years; (3) current use of 2 or more anti-seizure medications (ASMs) at stable doses; (4) average seizure frequency of 2 or more per 28 days; and (5) signed informed consent. Key exclusion criteria include (1) history of neurological or psychiatric disease (e.g., dementia, psychosis, or active suicidal ideation); (2) cerebrovascular history, including stroke or subarachnoid hemorrhage; (3) immunological risks (e.g., panel reactive antibody ≥ 20%, donor specific antibody positivity, or autoimmune disorders); (4) allergy or contraindication to immunosuppressants, prophylactic medications, or imaging reagents; (5) active malignancy or history of oncology related exclusions; and (6) pregnancy, breastfeeding, or any condition deemed unsuitable by the investigator.

### Rationale for Dose Selection

Intracranial cell products exhibit complex pharmacokinetic and pharmacodynamic profiles involving survival, proliferation, and synaptic integration. These factors preclude the use of conventional modeling to determine the minimum anticipated biological effect level (MABEL) or maximum tolerated dose (MTD). Consequently, dose selection is based on preclinical efficacy data, target cell density deficits in human hippocampal pathology and established safety margins.

#### Efficacy Dose Derivation

The primary therapeutic target is the restoration of somatostatin (SST) positive interneuron density within the hippocampus. Post-mortem data indicate that SST-positive neuron density in the hippocampus hilar region of healthy individuals is 655±127 neurons/mm^3^, whereas in patients with epileptogenic hippocampal sclerosis, density is reduced to 102±34 neurons/mm^3^.^26^ This difference represents a maximum deficit of approximately 646 neurons/mm^3^, providing the theoretical basis for functional restoration.

Preclinical studies in MTLE mouse models utilizing single intrahippocampal injections demonstrated that doses of 2.0×10^5^ (200K), and 1.5×10^6^ (1.5M) cells resulted in statistically significant seizure suppression at 7 months post-transplantation.^23^ Within the 2.5×10^4^ to 2.0×10^5^ cell dose range, human cell persistence averaged 7%–10% of the initial dose. However, the rate decreased to less than 2% at the 1.5M dose, suggesting a plateau in biological carrying capacity of the host hippocampus. Approximately 20%–30% of surviving cells differentiated into target functional SST positive neurons.

To derive the human effective doses, we estimated the functional SST positive neurons in mouse models (Table 2). At the 200K mouse dose, a conservative estimate of 2,800 surviving SST positive cells was established, and the estimate was 6,000 cells at the 1.5M dose. To achieve the target density of 646 SST+ neurons/mm^3^ in human hippocampus, we extrapolated the required concentration based on the mouse cellular conversion efficiencies: approximately 46,143 cells/µl (extrapolated from the 200K dose) and 161,500 cells/µl (extrapolated from the 1.5M dose). Given a planned total injection volume of 30 µl per human hippocampus, the final human doses are approximately 1.4 × 10^6^ cells for the low-dose group and 5.0 × 10^6^ cells for the high-dose group.

#### Safety Margin Assessment

In the NOG mouse model, a dose of 1.5 × 10^6^ cells/animal (approximately 3.1 × 10^9^ cells/kg brain weight) was determined as the No Observed Adverse Effect Level (NOAEL). No apparent systemic or target organ toxicities were observed at this dose.

The proposed human high dose of 5.0 × 10^6^ cells per hippocampus translates to approximately 3.76 × 10^6^ cells/kg brain weight for males and 4.20 × 10^6^ cells/kg brain weight for females based on standard human brain mass. Compared to the preclinical NOAEL, this clinical dose provides a safety margin exceeding 700 fold at the toxicological level (Table 3). When compared to the mouse pharmacodynamic effective dose (2.0 × 10^5^ cells/animal, equivalent to 0.41 × 10^9^ cells/kg brain weight), the human high dose maintains a safety margin of 90 to 100 fold.

This magnitude of safety margin is significantly higher than the standard conversion factors typically used for conventional drugs. This conservative strategy is designed to mitigate uncertainties related to species discrepancies, hippocampal region sensitivity, and the inherent complexity of iPSC-derived cell products.

### Recruitment and Consent

Potential participants are identified and recruited at the research site by investigators in the functional neurosurgery outpatient clinic. A study team member visits all patients meeting the eligibility criteria to invite them to participate in the study. For eligible individuals, written informed consent is obtained in the outpatient setting prior to enrollment and any study related procedures. The trial protocol and all recruitment materials have been approved by the institutional review board at the participating center.

### Randomization and Blinding

Participants are randomly assigned to the low dose or high dose group in a 1:1 ratio using a block randomization sequence. An independent biostatistician not involved in data collection or analysis generates the randomization list using SAS Version 9.4 (SAS Institute Inc.). Randomization sequence concealment is achieved through the randomization module of the Electronic Data Capture (EDC) system (Viedoc^TM)^. This system generates a unique randomization number for each participant and assigns a corresponding medication kit number for the investigator to administer.

To mitigate risk, a staggered enrollment strategy is utilized (Figure 1). Within each dose cohort, the first participant is monitored for at least 3 months following ALC05 injection. Enrollment of the remaining participants proceeds only after the initial participant demonstrates acceptable safety and tolerability, as reviewed by the Independent Data Safety Monitoring Board

Participants, investigators, and outcome assessors remain blinded to treatment allocation. Unblinding is permissible only if necessary for safety reasons.

### Intervention and Study Visits

#### Screening Visit

Following written informed consent, demographic data and medical history are obtained. Seizures are reviewed to calculate seizure frequency, and peripheral venous blood will be drawn for donor specific antibody and panel reactive antibody testing. Based on these preliminary results, investigators perform an initial screening to determine if participants tentatively meet the eligibility criteria.

#### Baseline Visit

Baseline assessments are conducted in the inpatient setting. For participants meeting initial screening requirements, comprehensive clinical evaluations are performed during the baseline period. All laboratory tests and scale assessments are completed according to the study schedule (Table 1). The investigator team confirms final enrollment based on the cumulative results from screening and baseline examinations.

**Table 1:**
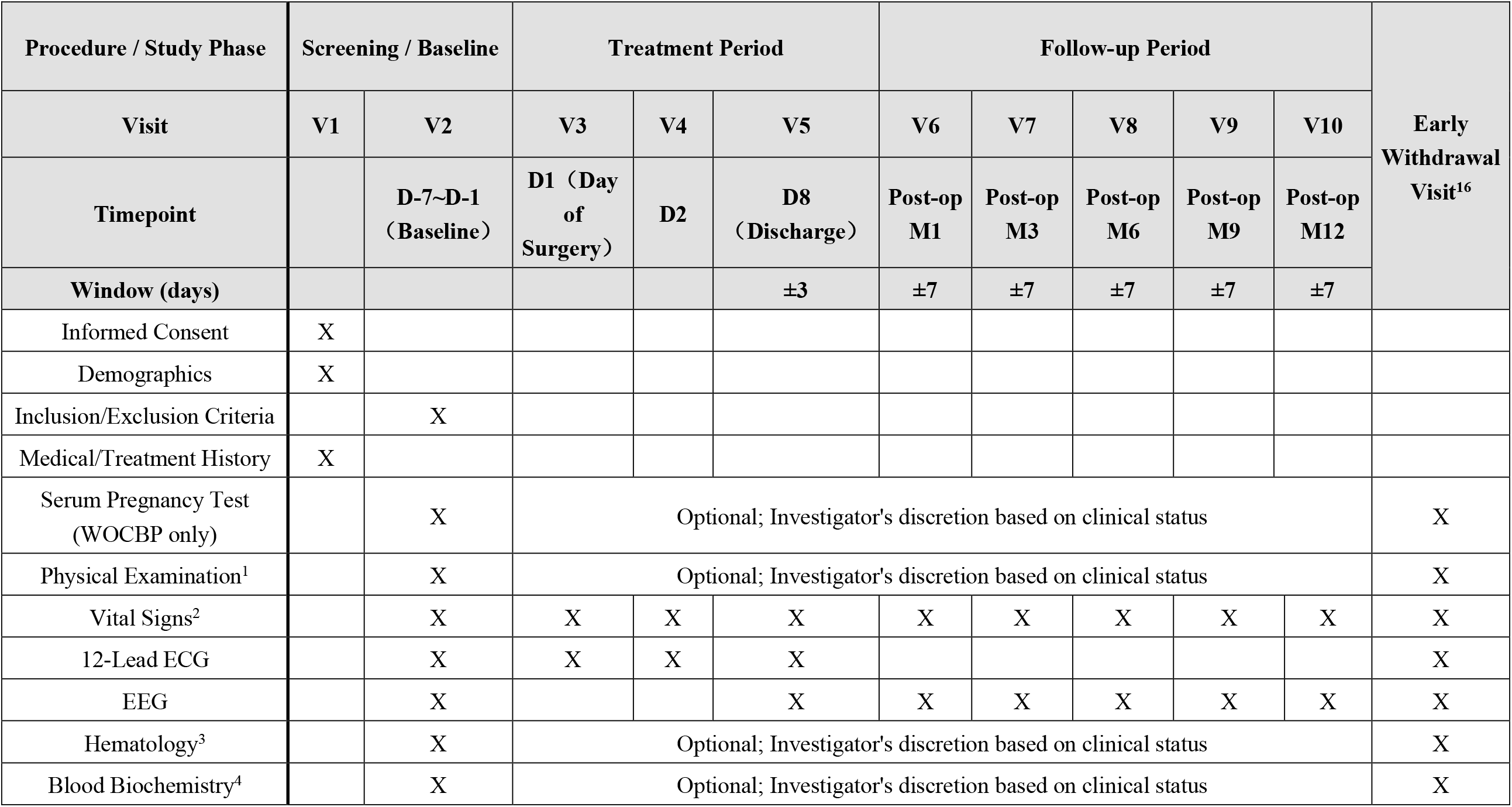
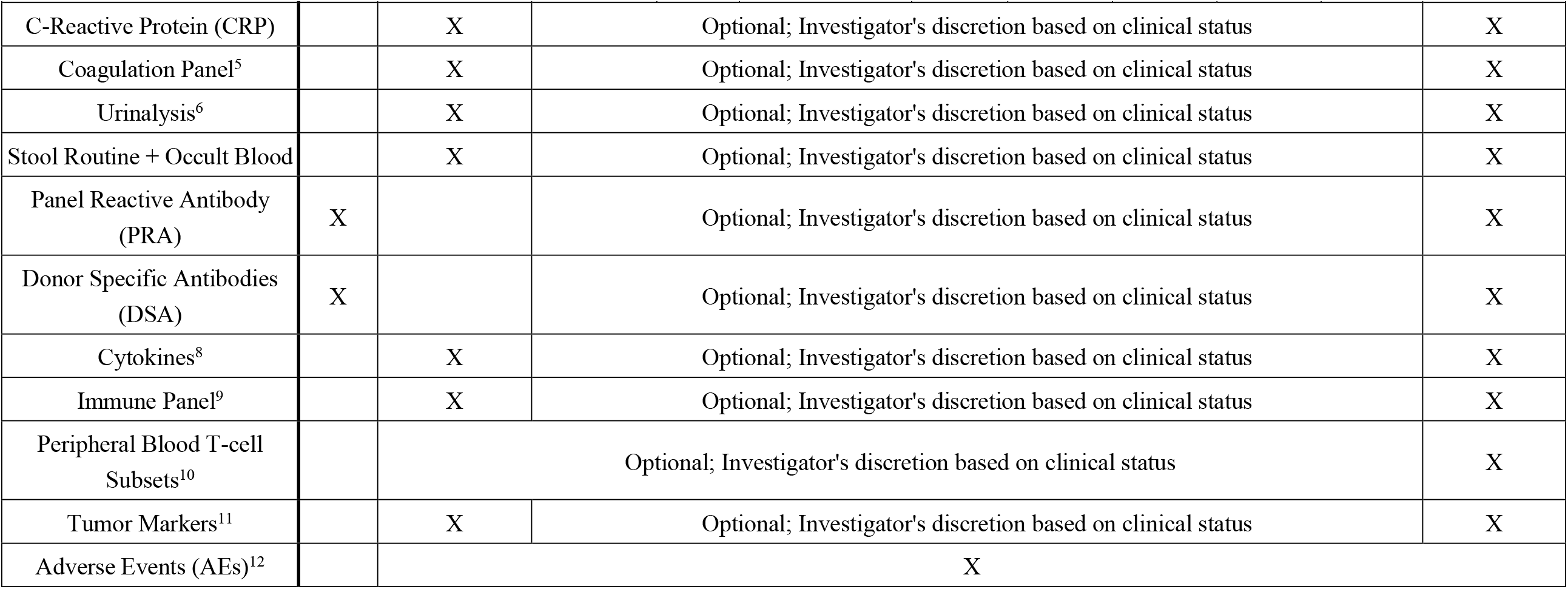
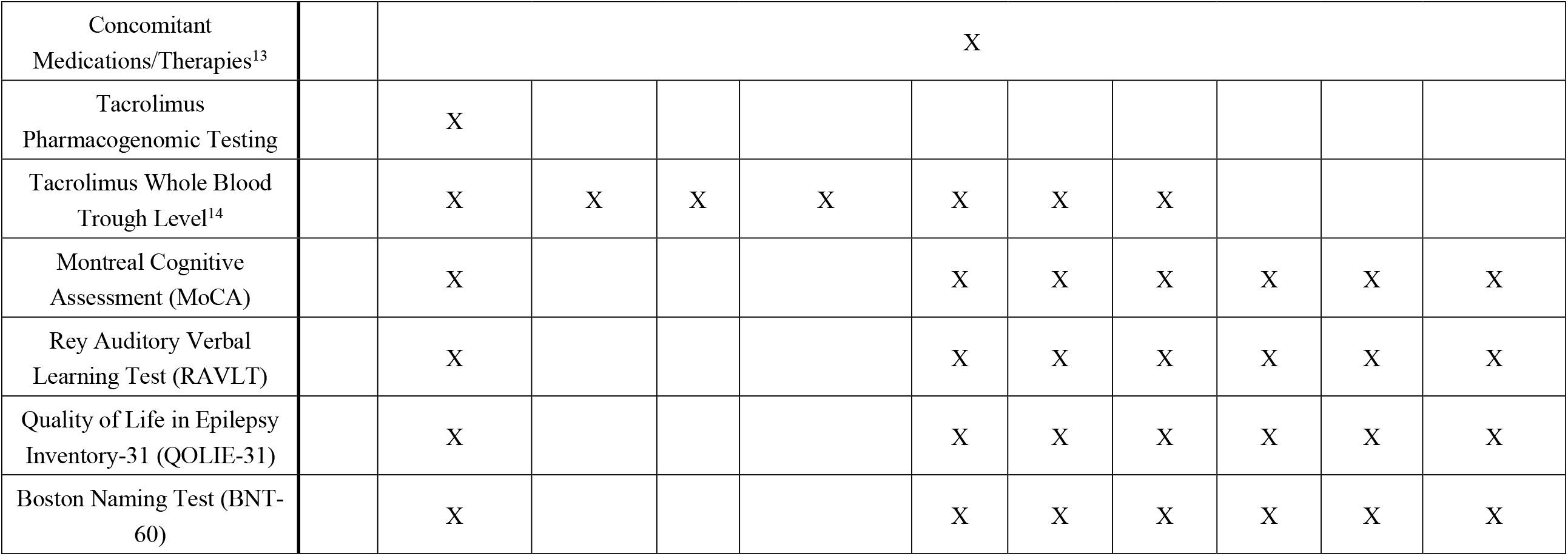
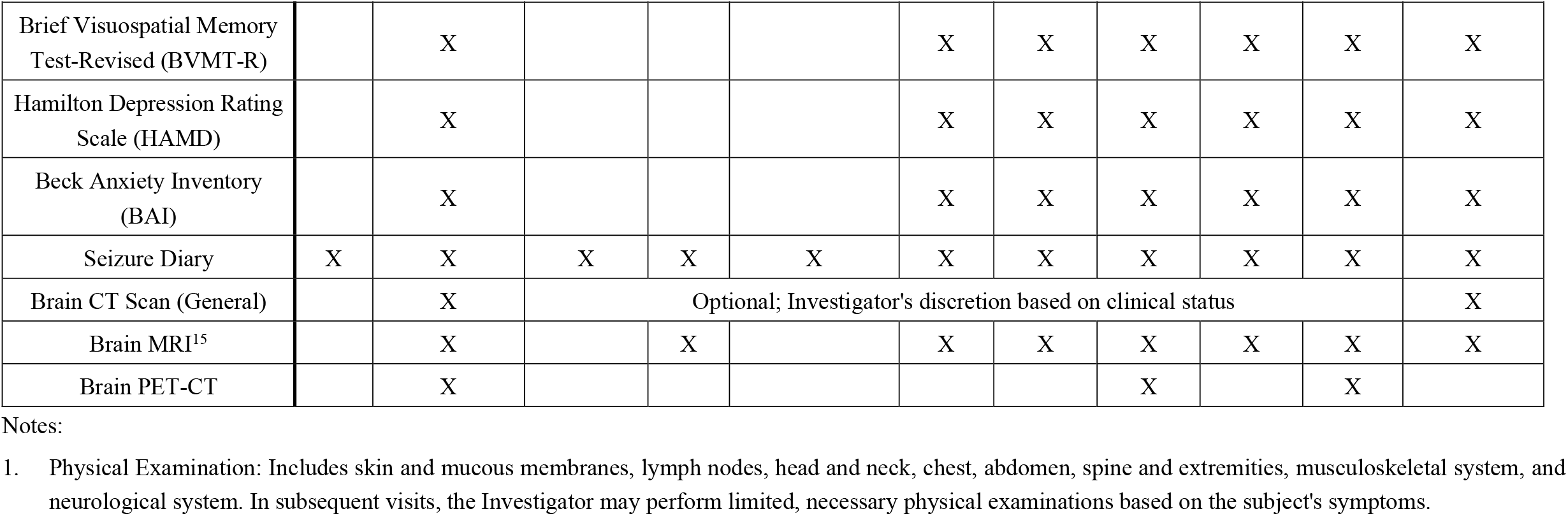
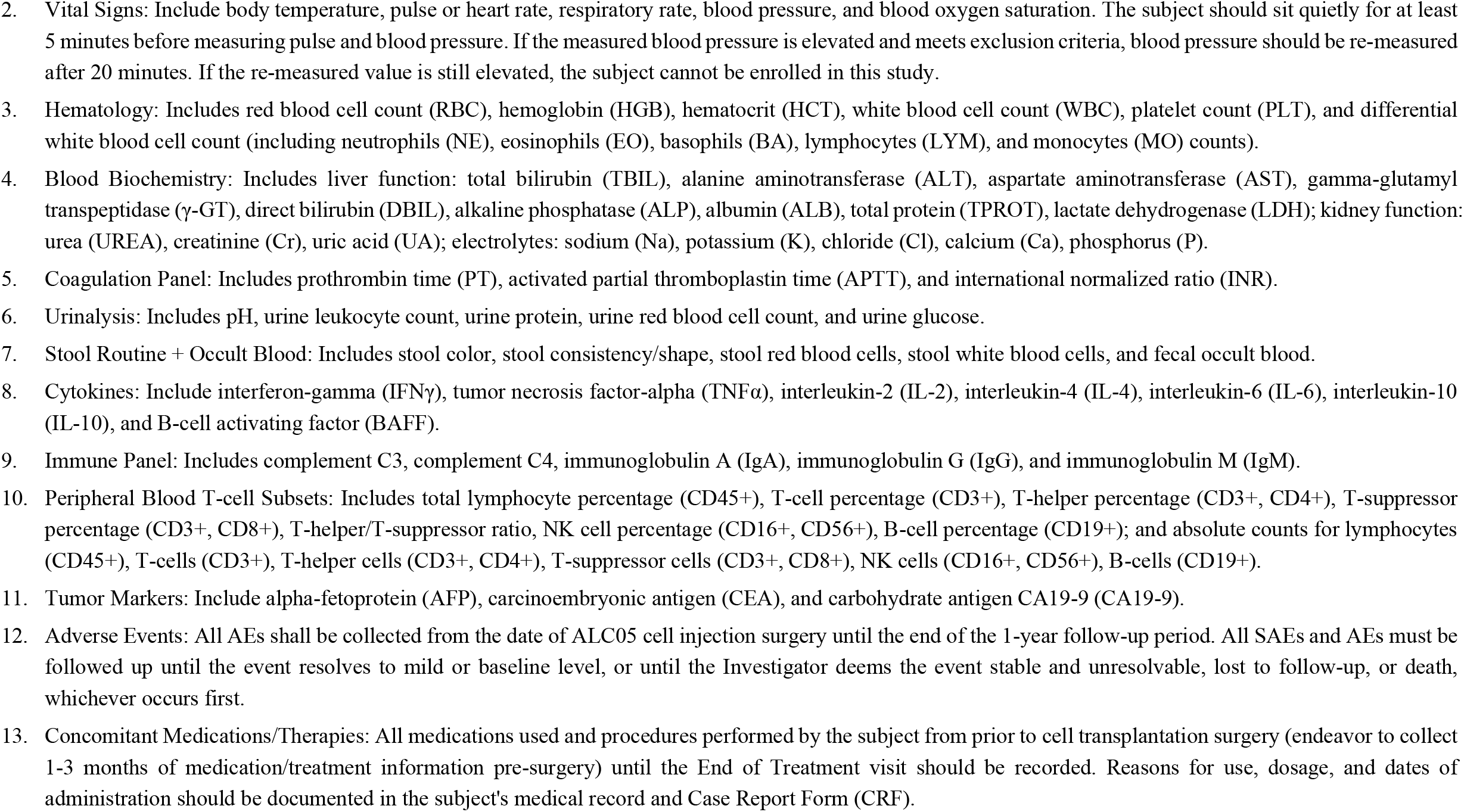
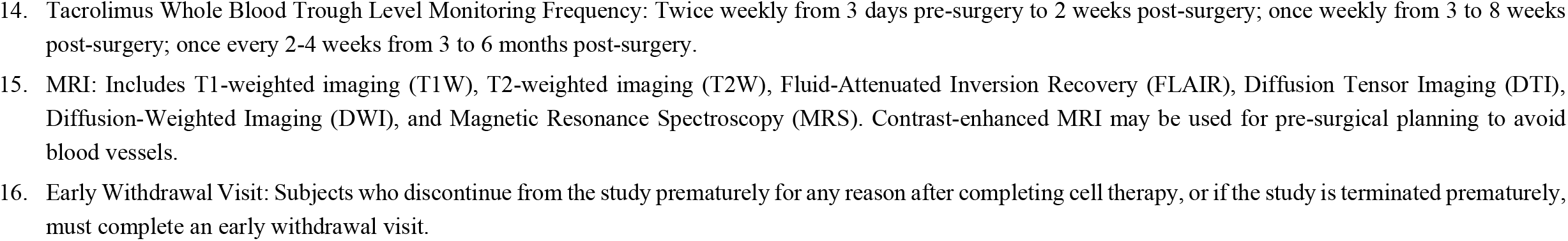
Study Schedule.

**Table 2:**
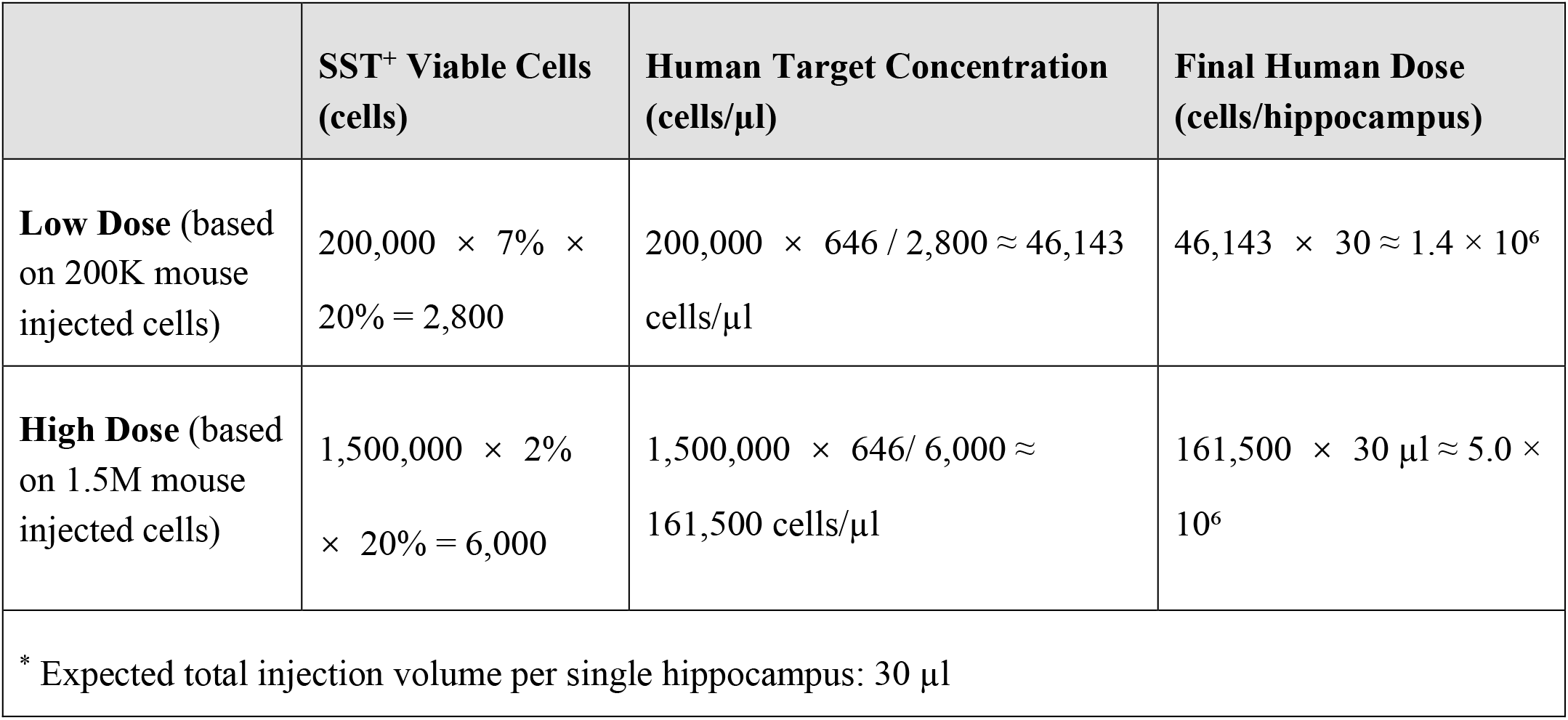
Derivation of Proposed Human Doses for SST+ Viable Interneuron Transplantation.

|  | **SST<sup>+</sup> Viable Cells**<br>**(cells)** | **Human Target Concentration**<br>**(cells/μl)** | **Final Human Dose**<br>**(cells/hippocampus)** |
| --- | --- | --- | --- |
| **Low Dose** (based on 200K mouse injected cells) | $200,000 \times 7\% \times 20\% = 2,800$ | $200,000 \times 646 / 2,800 \approx 46,143$<br>cells/μl | $46,143 \times 30 \approx 1.4 \times 10^6$ |
| **High Dose** (based on 1.5M mouse injected cells) | $1,500,000 \times 2\% \times 20\% = 6,000$ | $1,500,000 \times 646 / 6,000 \approx 161,500$<br>cells/μl | $161,500 \times 30 \mu\text{l} \approx 5.0 \times 10^6$ |
| * Expected total injection volume per single hippocampus: 30 μl | | | |

**Table 3:**
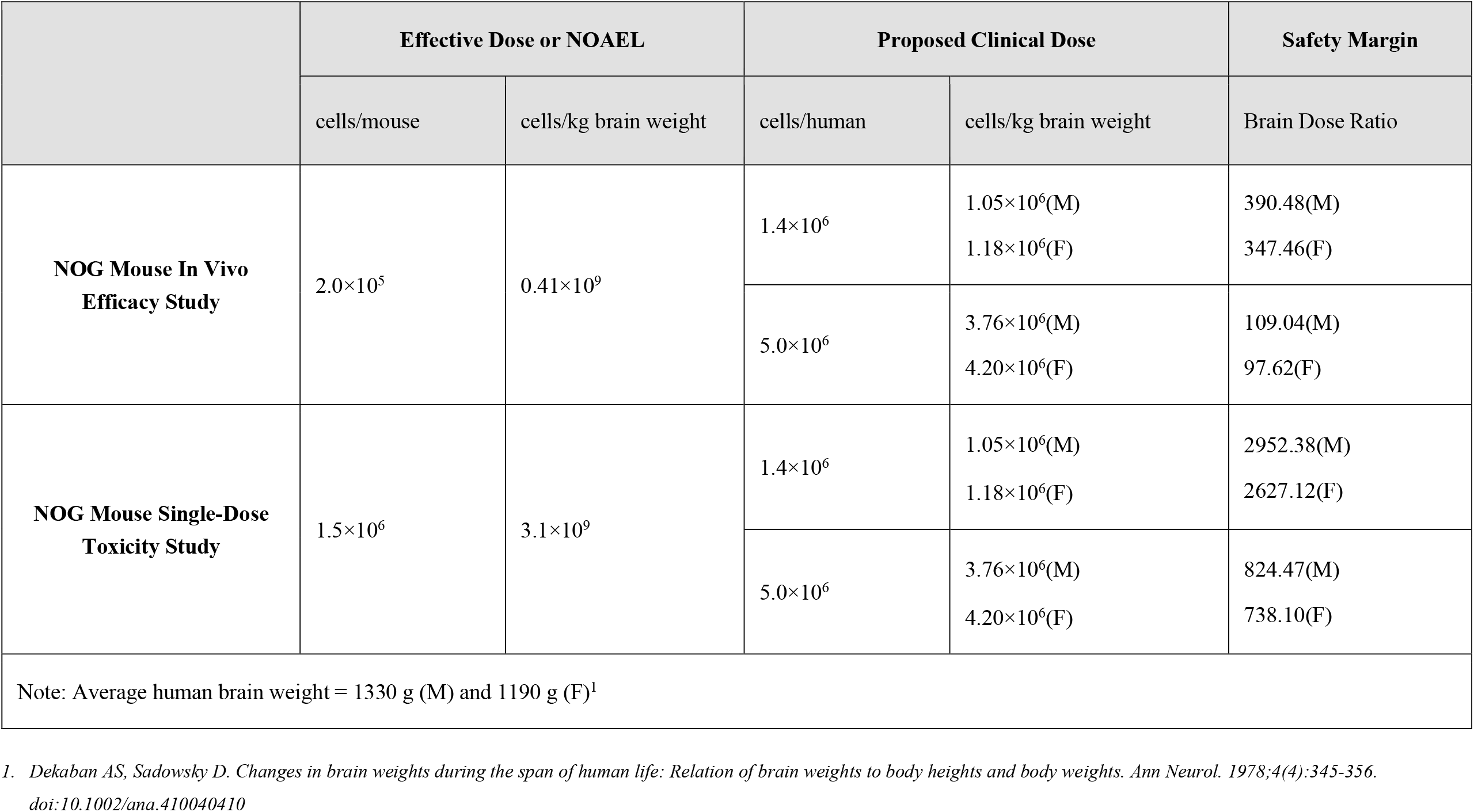
Preclinical to Clinical Dose Translation and Safety Margin Assessment.

#### Intervention

Following randomization, participants undergo standard preoperative monitoring, including electrocardiography, blood pressure, and pulse oximetry. Under general anesthesia, participants receive a stereotactic intracerebral injection of ALC05 cell suspension at the concentration corresponding to the assigned dose group. Vital signs and neurological status are monitored continuously throughout the perioperative period. Supportive care is provided as needed.

A comprehensive emergency preparedness plan has been established to manage potential complications. Postoperative imaging including MRI or CT of the transplant site is performed to assess complications such as hematoma. All clinical evaluations and laboratory tests are documented according to the study schedule (Table 1)

#### Follow-up Visits

Postoperative assessments are conducted on Day 7 post-transplantation. The primary follow-up period spans 12 months, with scheduled in person visits at Months 1, 3, 6, 9, and 12. Evaluation metrics during these visits include seizure frequency calculation, clinical scale assessments, and imaging examinations. Optional peripheral venous blood collection for laboratory tests is offered at each interval.

Upon completion of the 1-year study period, participants enter a long term safety surveillance phase until death or loss to follow-up. Subsequent evaluations occur at years 1, 2, 3, 5, 8, and 10 post-treatment via outpatient clinic visits. Follow up metrics for these long-term visits include seizure frequency, responder rates, and imaging examinations. Following the 10-year visit, participants are monitored every 5 years via telephone to determine survival status.

### Data Management

Data are collected through clinical medical records and scheduled participant visits. All assessments used for primary, secondary, and exploratory outcomes are performed at each scheduled interval (Table 1). A study specific electronic data set is generated using the Viedoc system. Consent forms and case report forms are secured in a research cabinet accessible only to study researchers and are retained for at least 15 years following trial completion at the study site.

Quality control procedures are applied throughout the study duration. To address uncertainty regarding the timing of events for participants who miss visits, sensitivity analyses are performed. For occurrences detected following one or more missed visits, the timing is set first to the detection date and second to the date of the missed visit prior to detection. Protocol violations, including failure to complete the full follow up schedule, are handled by comparison of intention to treat and per protocol analyses. All data are merged at the study conclusion and exported for further statistical analysis.

### Statistical Analysis

Statistical analyses are performed using SAS® Version 9.4 (Comp). All formal comparisons utilize two-sided tests with the significance level set at P<0.05. Given the exploratory nature of the first in human study, no adjustments for multiplicity are applied. Results are primarily intended for hypotheses generation and to guide future research.

#### Analysis Populations

The study uses three analysis populations. The full analysis set (FAS) includes all randomized participants who received the ALC05 treatment. This population serves as the primary basis for demographic characteristics, baseline features, and efficacy analysis. The safety analysis set comprises all participants who received ALC05 and had at least one post-baseline safety assessment. The per protocol set (PPS) consists of FAS participants who strictly adhered to the protocol, including an 80% procedure compliance rate and no major deviations. Efficacy analyses are conducted on both FAS and PPS.

#### Handling of Missing Data

No imputation is performed for missing demographic data or baseline characteristics. To address uncertainty arising from missing visits, sensitivity analyses are performed. For efficacy events detected following one or more missed visits, the event timing is set first to the detection date second to the date of the missed visit prior to detection. Additional sensitivity analyses for seizure frequency use the last observation carried forward (LOCF) method.

#### Outcome Analysis

The primary safety outcome is the incidence of treatment emergent adverse events. Secondary efficacy outcomes, including responder rates and changes in seizure frequency, are summarized by dose group. Continuous variables are presented as mean (SD) or median (interquartile range) based on distribution, and categorical variables are presented as numbers and percentages. For longitudinal outcomes, an observed case approach is used at each scheduled interval.

### Outcomes and Measures

#### Primary Outcome

The primary endpoint is the frequency and severity of treatment emergent adverse events occurring through 12 months post-transplantation. Safety assessments include physical and neurological examinations, vital signs, clinical laboratory evaluations, and the identification of adverse events of special interest, such as cytokine release syndrome and immune effector cell associated neurotoxicity syndrome.

#### Secondary Outcome

Secondary endpoints evaluate ALC05 biological activity and clinical seizure control through several measures. The responder rate is defined as the percentage of participants achieving a 50% or greater reduction in the frequency of focal seizures per 28 days compared with the pre-transplantation baseline. Seizure frequency is quantified by the change in the number of all focal seizures, with specific analysis of focal impaired awareness seizures and focal to bilateral tonic-clonic seizures.

Cell engraftment and survival are assessed via metabolic and functional changes at injection site using fluorodeoxyglucose positron emission tomography (FDG-PET) and magnetic resonance spectroscopy (MRS).

#### Exploratory Outcome

Exploratory endpoints evaluate the broader clinical and biological impact of ALC05 treatment. Patient-reported quality of life is assessed with the Quality of Life in Epilepsy-31 (QOLIE-31). Neurocognitive evaluation includes the Montreal Cognitive Assessment (MoCA) for global cognition, the Rey Auditory Verbal Learning Test (RAVLT) for verbal learning and memory, the Boston Naming Test (BNT-60) for language and naming, and the Brief Visuospatial Memory Test-Revised (BVMT-R) for visuospatial recall.

Psychological health is monitored through changes in mood and anxiety levels using the Hamilton Depression Rating Scale (HAMD) and the Beck Anxiety Inventory (BAI). The study also investigates potential associations between clinical seizure reduction and biological markers of graft activity, including metabolic changes on FDG PET and GABA concentration on MRS. All exploratory measures are expressed as changes from the pre-transplantation baseline.

## Discussion

For patients with drug-resistant MTLE, current treatment options have notable limitations. ASMs fail to control seizures in approximately 20% to 30% cases,^27^ and respective surgery carries risks of permanent neurological sequelae.^9^ Stem cell therapy has drawn interest as an alternative approach to addressing these limitations. This trial evaluates iPSC-derived GABAergic interneuron transplantation to restore inhibitory circuit function. Success in this trial would provide a therapeutic alternative for patients who are not suitable candidates for traditional surgical intervention.

Determining the optimal dose remains a primary challenge in the translation of regenerative therapies. Standard pharmacokinetic and pharmacodynamic modeling cannot fully account for survival, differentiation, and synaptic integration of transplanted cells. Dose selection in functional cell replacement therapy therefore relies on preclinical toxicology data. This study proposes a semi-quantitative dose derivation approach based on specific biomarkers and the estimated functional cellular deficits. This method supports biologically rational dose estimation, improving safety and optimizing cell use. It represents a shift toward more precise experimental design in cell therapy trials.

## Data Availability

All data produced in the present study are available upon reasonable request to the authors

## Limitations

Several limitations should be considered. The single-center design and small sample size (n=12) constrain the statistical power to detect subtle treatment effects. Thus, the findings may not be fully generalizable to broader MTLE population. Although the protocol includes a long term safety monitoring plan extending to 15 years, the primary observation period of 12 months may not sufficient to characterize the durability of graft function or to identify late-onset adverse events related to cellular integration and functional maturation.

## Conclusions

This Phase I randomized, double-blind clinical trial is designed to evaluate the safety and preliminary efficacy of ALC05, an allogeneic iPSC-derived GABAergic interneuron product, in patients with unilateral drug-resistant MTLE. The study addresses a clinical need that remains by current pharmacologic or surgical options. Results from this trial will provide initial safety data and contribute to the development of a biomarker-informed dosing framework in regenerative neurology.

## Article Information

### Author Contributions

J.Z. and B.T. had full access to all of the data in the study and take responsibility for the integrity of the data and the accuracy of the data analysis.

Concept and design: J.Z., B.T.

Acquisition, analysis, or interpretation of data: B.T.

Drafting of the manuscript: B.T.

Critical revision of the manuscript for important intellectual content: J.Z. Statistical analysis: J.Z.

Administrative, technical, or material support: J.Z. Supervision: J.Z.

