## supplement for "Safety and Efficacy of iPSC-Derived GABAergic Interneurons for Unilateral MTLE"

Study Protocol

#### LIST OF TABLES

Table 1 Study Schedule ........................................ 14

#### LIST OF FIGURES

Figure 1 Flow Diagram of the Trial ........................................ 30

### 1 Summary

<table>
<tr><td>Study Title</td><td>Safety and Efficacy of Human iPSC-Derived GABAergic Interneurons for the Treatment of Unilateral Mesial Temporal Lobe Epilepsy</td></tr>
<tr><td>Phase</td><td>Phase 1</td></tr>
<tr><td>Study Site</td><td>The First Affiliated Hospital of Xi'an Jiao Tong University</td></tr>
<tr><td>Sponsor</td><td>iCamuno Biotherapeutics Ltd.</td></tr>
<tr><td>Study Period</td><td>1 year</td></tr>
<tr><td>Studied Indication</td><td>Patients with Unilateral Mesial Temporal Lobe Epilepsy (MTLE)</td></tr>
<tr><td>Objectives</td><td>To explore the safety and preliminary efficacy of single-dose intracranial injection of ALC05 for the treatment of unilateral drug-resistant temporal lobe epilepsy, and to evaluate the therapeutic effects of different doses of ALC05 injection</td></tr>
<tr><td>Sample Size</td><td>The study involves two dose cohorts and plans to enroll a total of 12 subjects.
<table>
<tr><td></td><td>Sample Size</td></tr>
<tr><td>Low dose group</td><td>6 subjects</td></tr>
<tr><td>High dose group</td><td>6 subjects</td></tr>
</table></td></tr>
<tr><td>Study Design</td><td>Single-center, Randomized, Double-blind Phase 1 Clinical Trial</td></tr>
<tr><td>Study Method</td><td>This is a single-center, randomized, double-blind, phase 1 clinical trial designed to evaluate the safety and preliminary efficacy of ALC05 injection, a preparation of gene-edited, hypoimmunogenic iPSC-derived GABAergic interneurons.<br><br>The study plans to enroll 12 subjects with unilateral mesial temporal lobe epilepsy (MTLE) to receive stereotactic intracranial injections of ALC05. All enrolled subjects will be randomly divided into either low dose or high dose group at a 1:1 ratio. To minimize risks to subjects at each dose level, the first subject in each dose group will be monitored for safety for at least 3 months post ALC05 injection and show acceptable safety and tolerability, before the remaining patients can be enrolled. The</td></tr>
</table>

| | |
|---|---|
| | safety data will be reviewed by the Independent Data Safety Monitoring Board (DSMB).<br><br>**Study Procedures：**<br><br>Screening and Baseline Assessment: After obtaining informed consent, participants will undergo an initial screening to collect demographic and medical history, seizure diaries, and peripheral venous blood for DSA and PRA testing. Those tentatively meeting inclusion criteria will proceed to a baseline visit in the inpatient ward for a more comprehensive assessment, including detailed laboratory tests and standardized scale assessments. Final enrollment will be confirmed based on the complete results from both screening and baseline periods.<br><br>Intervention (ALC05 Cell Transplantation): Following randomization, participants will undergo standard preoperative monitoring. Upon confirmation of surgical eligibility, stereotactic intracranial injection of ALC05 cell suspension (either low- or high-dose group) will be performed under general anesthesia. Throughout the perioperative period, participants will receive close monitoring of vital signs and neurological status, supportive care, and management of potential complications as outlined in the emergency preparedness plan. Postoperative imaging (MRI or CT) will assess the transplant site for complications like hematoma.<br><br>Follow-up: A postoperative assessment will be conducted on Day 7 post-transplantation. The primary study follow-up period is 1 year, with in-person visits at Months 1, 3, 6, 9, and 12 for seizure frequency calculation, scale assessments, imaging examinations, and optional laboratory tests. After this 1-year period, participants will enter a long-term post-study safety follow-up phase until death or loss to follow-up. |
| Inclusion/ Exclusion Criteria | **[Inclusion Criteria]**<br><br>Subjects must meet all of the following criteria.<br><br>1. Capable of signing the informed consent form and complying with the study protocol.<br>2. Male or female, aged 18 to 75 years (inclusive).<br>3. Mesial Temporal Lobe Epilepsy (MTLE) confirmed by long-term scalp electroencephalography (EEG) monitoring, cranial imaging, and clinical symptoms.<br>4. Currently receiving adequate doses, sufficient duration, and rational use of ⩾2 anti-seizure medications (ASMs) approved by the National Medical |

Products Administration (NMPA), with a treatment duration of ⩾1 month, yet failing to achieve seizure control.

5. Average seizure frequency of ≥ 2 seizures per 28 days during the 6 months prior to the screening visit.
6. Deemed by the investigator to be a suitable candidate for Temporal Lobe Resection (TLR) or Laser Interstitial Thermal Therapy (LITT).

**[Exclusion Criteria]**

Subjects who meet any of the following criteria cannot participate in this study.

1. Patients with any of the following co-existing conditions: a) Other significant brain structural abnormalities (e.g., brain tumors, vascular malformations, arachnoid cysts, tuberous sclerosis, cortical dysplasia, etc.); b) History of Status Epilepticus within 1 year prior to screening (based on ILAE criteria, Trinka 2015, as judged by the Principal Investigator; however, a history of cluster seizures is permitted); c) Autoimmune epilepsy, with relevant serum antibodies detected within 3 years prior to screening or at baseline (accepted methods include Labcorp 505490, Mayo Clinic EPS2, or similar tests approved by the Sponsor); d) Progressive neurological diseases (e.g., multiple sclerosis, primary mitochondrial diseases, other neurodegenerative diseases); e) History of severe cranial diseases (e.g., stroke, traumatic brain injury, subarachnoid hemorrhage); f) Severe cognitive impairment, intellectual disability, or psychiatric disorders (e.g., schizophrenia, severe depression) that prevent completion of study procedures or provision of valid informed consent; g) History of suicidal behavior or suicidal ideation within 12 months prior to enrollment;
2. Unstable vital signs at screening and/or prior to surgery: a. Heart rate < 50 beats per minute (bpm) or > 105 bpm; b. Systolic Blood Pressure (SBP) < 90 mmHg or > 160 mmHg; Diastolic Blood Pressure (DBP) < 60 mmHg or > 100 mmHg; c. Respiratory rate < 12 breaths per minute or > 20 breaths per minute; d. Resting pulse oxygen saturation (SpO2) < 94%; e. Temperature > 100.4°F (38.0°C);
3. Estimated Glomerular Filtration Rate (eGFR) < 30 ml/min/1.73$m^2$, which cannot be corrected;
4. Abnormal liver function: ALT or AST > 3 times the upper limit of normal (ULN), which cannot be corrected;
5. Hematological abnormalities: e.g., Hematocrit < 25%, White Blood Cell count < 2,500/μL, or Platelet count < 100,000/μL, which cannot be corrected;

| | |
|---|---|
| | 6. Coagulation abnormalities: INR > 1.3, which cannot be corrected;<br>7. Patients with autoimmune diseases;<br>8. Long-term use of immunosuppressants, such as systemic corticosteroids, TNF-α inhibitors, etc.;<br>9. History of prior organ transplantation;<br>10. Patients with diabetes mellitus (high risk of underlying disease worsening or new infections);<br>11. Positive for Human Immunodeficiency Virus (HIV), Hepatitis B Virus (HBV), or Hepatitis C Virus (HCV);<br>12. Positive Panel Reactive Antibody (PRA);<br>13. Patients who have received cell or gene therapy (autologous or allogeneic);<br>14. Prior non-pharmacological treatments (e.g., surgical resection, thermal ablation, radiation therapy, VNS, DBS, RNS);<br>15. Patients participating in investigational drug or device trials at the time of signing informed consent or within 30 days after obtaining informed consent;<br>16. Patients unable to undergo MRI and PET-CT examinations;<br>17. Patients allergic to radiocontrast agents who cannot be managed with premedication;<br>18. History of allergy to methotrexate, ruxolitinib, or penicillin, or non-sensitive to tacrolimus based on genetic testing;<br>19. Patients with progressive malignant tumors within the past 5 years (excluding basal cell carcinoma);<br>20. Patients with a life expectancy of < 1 year;<br>21. Pregnant or breastfeeding women;<br>22. Women of Childbearing Potential (WOCBP) who cannot commit to strict adherence to effective contraception;<br>23. Any other patients deemed unsuitable for participation in this clinical study by the investigator. |
| Criteria for Withdrawal | **1. Withdrawal at the Investigator's Discretion**<br>This refers to instances where the Investigator decides that an enrolled subject is no longer suitable to continue in the study.<br>1) Poor subject compliance that affects the assessment of safety and tolerability. |

| | |
|---|---|
| | 2) Occurrence of an adverse event that, upon analysis, makes the subject unsuitable to continue in the study.<br>3) Other circumstances where the Investigator deems the subject unsuitable to continue in the study.<br>**2. Withdrawal by the Subject**<br>1) Subjects have the right to withdraw from the study at any time for any reason.<br>2) If a subject does not explicitly request to withdraw but fails to attend visits, receive the cell product transplantation, or cooperate with testing as required by the protocol, and the Investigator is unable to contact the subject on at least three occasions between the current and subsequent visit windows, the subject may be considered "lost to follow-up." This type of loss to follow-up is also classified as "withdrawal". |
| Duration, and Method of Administration | **Duration of administration:** Single administration<br>**Transplantation Sites:** Hippocampus |
| Permitted Concomitant Medications and Prohibited/Restricted Medications | **Permitted Concomitant Medications**<br>For medications taken prior to signing the Informed Consent Form (ICF), the current regimen may be maintained. For new symptoms or conditions emerging during the study, medications may be administered based on the judgment of the Investigator or designee, including but not limited to the following situations:<br>1) Use of Anti-Seizure Medications (ASMs): The use, dosage, and administration of other ASMs will be determined by the Investigator based on the subject's condition.<br>2) Supportive Therapy: The Investigator is permitted to use supportive therapies in accordance with the clinical site's medical guidelines. These include, but are not limited to, antibiotics, platelet transfusions, plasma transfusions, bronchodilators, epinephrine, antihistamines, intracranial decompression drugs, or glucocorticoids.<br>3) This study will develop personalized immunosuppressive strategies based on the participants' tacrolimus genetic testing results.<br>**Prohibited/Restricted Medications**<br>In general, any concomitant medication/treatment required by the subject is permitted, with the exception of the following:<br>1) Prohibited Epilepsy Treatments During the Study: Subjects are prohibited from receiving the following treatments during the study period, including |

| | |
|---|---|
| | but not limited to Temporal Lobe Resection (TLR), Laser Interstitial Thermal Therapy (LITT), other autologous/allogeneic cell transplantation/infusion, or participation in other clinical studies indicated for epilepsy.<br>2) Prohibition on Other Studies: Participation in other clinical studies involving cells, drugs, or devices is prohibited for 2 years post-surgery. |
| Endpoints | **Primary Endpoint:**<br>1. The frequency and severity of Adverse Events (AEs) and Serious Adverse Events (SAEs) within 12 months post-surgery.<br>**Secondary Endpoints:**<br>Efficacy:<br>1. Cell Engraftment and Survival: Changes in the metabolic and functional status of brain tissue assessed by Fluorodeoxyglucose Positron Emission Tomography (FDG-PET) and Magnetic Resonance Spectroscopy (MRS) compared to baseline (pre-transplantation).<br>2. Responder Rate: The percentage of subjects with a ≥50% reduction in seizure frequency compared to baseline.<br>3. Seizure Frequency: Change in seizure frequency (number of seizures per 28 days) compared to baseline; including the frequency of all seizure events and disabling seizures.<br>**Exploratory Endpoints:**<br>1. Quality of Life Assessment: Change in the Quality of Life in Epilepsy Inventory-31 (QOLIE-31) scores compared to baseline.<br>2. Neurocognitive Function Assessment: Changes in scores (or results) compared to baseline for the following: Montreal Cognitive Assessment (MoCA), Rey Auditory Verbal Learning Test (RAVLT), Boston Naming Test (BNT-60), and Brief Visuospatial Memory Test-Revised (BVMT-R).<br>3. Mood Disorder Assessment: Changes in scores for the Hamilton Depression Rating Scale (HAMD) and Beck Anxiety Inventory (BAI) compared to baseline. |
| Analysis Method | **Analysis Datasets:**<br>• Full Analysis Set (FAS): All enrolled subjects who have received ALC05 treatment constitute the Full Analysis Set for this study. The FAS population |

| | |
|---|---|
| | will be used for the analysis of demographic data, baseline characteristics, and efficacy.<br>• Safety Set (SS/SAS): All enrolled subjects who have at least one post-baseline safety evaluation data constitute the Safety Set for this study. The Safety Set will be used for safety data analysis.<br>• Per-Protocol Set (PPS): Subjects who strictly followed the clinical trial protocol and completed all treatments and follow-ups constitute the Per-Protocol Set for this study.<br>**Sample Size Determination:**<br>This study plans to enroll 12 subjects. As this is an exploratory study, there are no formal statistical hypotheses. It is anticipated that enrolling 12 subjects will satisfy the exploratory needs of this study regarding preliminary safety and efficacy.<br>**Safety Analysis:**<br>Safety analysis will be performed on the Safety Set and analyzed according to the treatment actually received. Appropriate descriptive statistics will be used to summarize safety data. Baseline is defined as the last non-missing value prior to receiving the ALC05 surgical transplantation. Summaries will be provided for demographics, vital signs, physical examination, ECG, pulmonary function tests, laboratory tests, concomitant medications, Adverse Events (AEs), and Serious Adverse Events (SAEs). All values outside the clinical reference range will be flagged in data listings. The primary analysis will focus on Treatment-Emergent Adverse Events (TEAEs) highly related to the study drug. AEs and SAEs will be coded using the Medical Dictionary for Regulatory Activities (MedDRA) and summarized by System Organ Class (SOC) and severity. If the severity of the same AE changes within the same admission cycle for the same case but does not resolve to normal, it is recorded as a single AE event. The time (only date required) is recorded as the time of the first occurrence, and the severity is recorded as the most severe grade observed until the AE resolves to normal.<br>**Efficacy Assessment:**<br>The primary analysis population for all efficacy endpoints is the Full Analysis Set (FAS), which includes all randomized subjects and follows the Intention-to-Treat (ITT) principle. Sensitivity analysis will be performed using the Per-Protocol Set (PPS), which consists of subjects meeting key protocol criteria without major deviations. All efficacy analyses will be performed by treatment group and for the overall study population. For continuous endpoints, summary statistics (n, mean, standard deviation, median, minimum, maximum) will be provided for each planned post-baseline visit/time point, and changes from baseline for each visit will be |

| | |
|---|---|
| | calculated and summarized. For categorical endpoints, counts and percentages will be listed by treatment group.<br><br>Responder Rate: Descriptive summaries of response rates will include the number (n), percentage (%), and 95% Confidence Interval (CI) of responders, non-responders, and seizure-free subjects for each time period (M1-M3, M4-M6, M7-M9, M10-M12) and overall (M1-M12), summarized by treatment group. The 95% CI for percentages will be calculated using the Clopper-Pearson exact method. Inter-group comparisons will use Fisher's Exact Test (two-sided, $\alpha$=0.05) to test for differences in response rates (responder vs. non-responder) between treatment groups (Low-Dose vs. High-Dose). Odds Ratios (OR) and their associated 95% CIs will be reported.<br><br>Seizure Frequency: Descriptive statistics for seizure frequency: For each time point and treatment group, summary statistics will be calculated for absolute seizure frequency, change from baseline, and percentage change from baseline. Summary statistics will include: number of subjects (n), Mean ± Standard Deviation (SD), Median, First Quartile (Q1), Third Quartile (Q3), Minimum (Min), Maximum (Max), and the 95% Confidence Interval (CI) of the mean. Intra-group analysis will test whether changes in seizure frequency from baseline at each time point (M1 to M12) are statistically significant. A paired t-test will be used for normally distributed data; the Wilcoxon signed-rank test will be used for non-normally distributed data. P-values and 95% CIs for mean change will be reported. Inter-group comparison will analyze the difference in absolute change from baseline between the Low-Dose and High-Dose groups at each time point. An independent samples t-test will be used for normally distributed data, and the Wilcoxon rank-sum test will be used for non-normally distributed data. The mean difference, its 95% CI, and descriptive p-values will be reported.<br><br>Scale Analysis: Descriptive statistics for scale analysis include subscale scores and total scores. Scores at baseline and all post-baseline visits will be summarized by treatment group. Changes in scores from baseline will be calculated and summarized at each time point. Summary statistics will include: n, mean ± SD, median, Q1, Q3, Min, Max, and 95% CI. Responder analysis will define clinically meaningful improvement as an increase in total score from baseline of ≥X points (defined according to the Minimal Clinically Important Difference [MCID] for different scales). The number (n) and percentage (%) of subjects achieving this clinically meaningful improvement will be calculated, and associated 95% CIs determined using the Clopper-Pearson exact method. Intra-group analysis will use paired t-tests at each post-baseline visit to test for statistical differences in subscale and total score changes from baseline. Effect size (Cohen's d) will be calculated to assess the magnitude of efficacy. Inter-group comparison will use Analysis of Covariance (ANCOVA) models at each visit to compare the "change in score from baseline" |

| | |
|---|---|
| | between treatment groups: *Change from Baseline = Treatment Group + Baseline Score*. The Least Squares (LS) mean difference between groups, its 95% CI, and descriptive p-values will be reported. |

**Table 1 Study Schedule**

| Procedure / Study Phase | Screening / Baseline | | Treatment Period | | | Follow-up Period | | | | | Early Withdrawal Visit[16] |
|---|---|---|---|---|---|---|---|---|---|---|---|
| **Visit** | **V1** | **V2** | **V3** | **V4** | **V5** | **V6** | **V7** | **V8** | **V9** | **V10** | |
| **Timepoint** | | **D-7~D-1（Baseline）** | **D1（Day of Surgery）** | **D2** | **D8（Discharge）** | **Post-op M1** | **Post-op M3** | **Post-op M6** | **Post-op M9** | **Post-op M12** | |
| **Window (days)** | | | | | **±3** | **±7** | **±7** | **±7** | **±7** | **±7** | |
| Informed Consent | X | | | | | | | | | | |
| Demographics | X | | | | | | | | | | |
| Inclusion/Exclusion Criteria | | X | | | | | | | | | |
| Medical/Treatment History | X | | | | | | | | | | |
| Serum Pregnancy Test (WOCBP only) | | X | Optional; Investigator's discretion based on clinical status | | | | | | | | X |
| Physical Examination[1] | | X | Optional; Investigator's discretion based on clinical status | | | | | | | | X |
| Vital Signs[2] | | X | X | X | X | X | X | X | X | X | X |
| 12-Lead ECG | | X | X | X | X | | | | | | X |
| EEG | | X | | | X | X | X | X | X | X | X |
| Hematology[3] | | X | Optional; Investigator's discretion based on clinical status | | | | | | | | X |
| Blood Biochemistry[4] | | X | Optional; Investigator's discretion based on clinical status | | | | | | | | X |
| C-Reactive Protein (CRP) | | X | Optional; Investigator's discretion based on clinical status | | | | | | | | X |
| Coagulation Panel[5] | | X | Optional; Investigator's discretion based on clinical status | | | | | | | | X |
| Urinalysis[6] | | X | Optional; Investigator's discretion based on clinical status | | | | | | | | X |
| Stool Routine + Occult Blood | | X | Optional; Investigator's discretion based on clinical status | | | | | | | | X |

| Procedure / Study Phase | Screening / Baseline | | Treatment Period | | | Follow-up Period | | | | | Early Withdrawal Visit[16] |
|---|---|---|---|---|---|---|---|---|---|---|---|
| **Visit** | **V1** | **V2** | **V3** | **V4** | **V5** | **V6** | **V7** | **V8** | **V9** | **V10** | |
| **Timepoint** | | **D-7~D-1（Baseline）** | **D1（Day of Surgery）** | **D2** | **D8（Discharge）** | **Post-op M1** | **Post-op M3** | **Post-op M6** | **Post-op M9** | **Post-op M12** | |
| **Window (days)** | | | | | **±3** | **±7** | **±7** | **±7** | **±7** | **±7** | |
| Panel Reactive Antibody (PRA) | X | | Optional; Investigator's discretion based on clinical status | | | | | | | | X |
| Donor Specific Antibodies (DSA) | X | | Optional; Investigator's discretion based on clinical status | | | | | | | | X |
| Cytokines[8] | | X | Optional; Investigator's discretion based on clinical status | | | | | | | | X |
| Immune Panel[9] | | X | Optional; Investigator's discretion based on clinical status | | | | | | | | X |
| Peripheral Blood T-cell Subsets[10] | | Optional; Investigator's discretion based on clinical status | | | | | | | | | X |
| Tumor Markers[11] | | X | Optional; Investigator's discretion based on clinical status | | | | | | | | X |
| Adverse Events (AEs)[12] | | X | | | | | | | | | |
| Concomitant Medications/Therapies[13] | | X | | | | | | | | | |
| Tacrolimus Pharmacogenomic Testing | | X | | | | | | | | | |
| Tacrolimus Whole Blood Trough Level[14] | | X | X | X | X | X | X | X | | | |
| Montreal Cognitive Assessment (MoCA) | | X | | | | X | X | X | X | X | X |
| Rey Auditory Verbal Learning Test (RAVLT) | | X | | | | X | X | X | X | X | X |
| Quality of Life in Epilepsy Inventory-31 (QOLIE-31) | | X | | | | X | X | X | X | X | X |

| Procedure / Study Phase | Screening / Baseline | | Treatment Period | | | Follow-up Period | | | | | Early Withdrawal Visit[16] |
|---|---|---|---|---|---|---|---|---|---|---|---|
| **Visit** | **V1** | **V2** | **V3** | **V4** | **V5** | **V6** | **V7** | **V8** | **V9** | **V10** | |
| **Timepoint** | | **D-7~D-1（Baseline）** | **D1（Day of Surgery）** | **D2** | **D8（Discharge）** | **Post-op M1** | **Post-op M3** | **Post-op M6** | **Post-op M9** | **Post-op M12** | |
| **Window (days)** | | | | | **±3** | **±7** | **±7** | **±7** | **±7** | **±7** | |
| Boston Naming Test (BNT-60) | | X | | | | X | X | X | X | X | X |
| Brief Visuospatial Memory Test-Revised (BVMT-R) | | X | | | | X | X | X | X | X | X |
| Hamilton Depression Rating Scale (HAMD) | | X | | | | X | X | X | X | X | X |
| Beck Anxiety Inventory (BAI) | | X | | | | X | X | X | X | X | X |
| Seizure Diary | X | X | X | X | X | X | X | X | X | X | X |
| Brain CT Scan (General) | | X | Optional; Investigator's discretion based on clinical status | | | | | | | | X |
| Brain MRI[15] | | X | | X | | X | X | X | X | X | X |
| Brain PET-CT | | X | | | | | | X | | X | |

Notes:

1. Physical Examination: Includes skin and mucous membranes, lymph nodes, head and neck, chest, abdomen, spine and extremities, musculoskeletal system, and neurological system. In subsequent visits, the Investigator may perform limited, necessary physical examinations based on the subject's symptoms.
2. Vital Signs: Include body temperature, pulse or heart rate, respiratory rate, blood pressure, and blood oxygen saturation. The subject should sit quietly for at least 5 minutes before measuring pulse and blood pressure. If the measured blood pressure is elevated and meets exclusion criteria, blood pressure should be re-measured after 20 minutes. If the re-measured value is still elevated, the subject cannot be enrolled in this study.
3. Hematology: Includes red blood cell count (RBC), hemoglobin (HGB), hematocrit (HCT), white blood cell count (WBC), platelet count (PLT), and differential white blood cell count (including neutrophils (NE), eosinophils (EO), basophils (BA), lymphocytes (LYM), and monocytes (MO) counts).

4. Blood Biochemistry: Includes liver function: total bilirubin (TBIL), alanine aminotransferase (ALT), aspartate aminotransferase (AST), gamma-glutamyl transpeptidase (γ-GT), direct bilirubin (DBIL), alkaline phosphatase (ALP), albumin (ALB), total protein (TPROT), lactate dehydrogenase (LDH); kidney function: urea (UREA), creatinine (Cr), uric acid (UA); electrolytes: sodium (Na), potassium (K), chloride (Cl), calcium (Ca), phosphorus (P).
5. Coagulation Panel: Includes prothrombin time (PT), activated partial thromboplastin time (APTT), and international normalized ratio (INR).
6. Urinalysis: Includes pH, urine leukocyte count, urine protein, urine red blood cell count, and urine glucose.
7. Stool Routine + Occult Blood: Includes stool color, stool consistency/shape, stool red blood cells, stool white blood cells, and fecal occult blood.
8. Cytokines: Include interferon-gamma (IFNγ), tumor necrosis factor-alpha (TNFα), interleukin-2 (IL-2), interleukin-4 (IL-4), interleukin-6 (IL-6), interleukin-10 (IL-10), and B-cell activating factor (BAFF).
9. Immune Panel: Includes complement C3, complement C4, immunoglobulin A (IgA), immunoglobulin G (IgG), and immunoglobulin M (IgM).
10. Peripheral Blood T-cell Subsets: Includes total lymphocyte percentage (CD45+), T-cell percentage (CD3+), T-helper percentage (CD3+, CD4+), T-suppressor percentage (CD3+, CD8+), T-helper/T-suppressor ratio, NK cell percentage (CD16+, CD56+), B-cell percentage (CD19+); and absolute counts for lymphocytes (CD45+), T-cells (CD3+), T-helper cells (CD3+, CD4+), T-suppressor cells (CD3+, CD8+), NK cells (CD16+, CD56+), B-cells (CD19+).
11. Tumor Markers: Include alpha-fetoprotein (AFP), carcinoembryonic antigen (CEA), and carbohydrate antigen CA19-9 (CA19-9).
12. Adverse Events: All AEs shall be collected from the date of ALC05 cell injection surgery until the end of the 1-year follow-up period. All SAEs and AEs must be followed up until the event resolves to mild or baseline level, or until the Investigator deems the event stable and unresolvable, lost to follow-up, or death, whichever occurs first.
13. Concomitant Medications/Therapies: All medications used and procedures performed by the subject from prior to cell transplantation surgery (endeavor to collect 1-3 months of medication/treatment information pre-surgery) until the End of Treatment visit should be recorded. Reasons for use, dosage, and dates of administration should be documented in the subject's medical record and Case Report Form (CRF).
14. Tacrolimus Whole Blood Trough Level Monitoring Frequency: Twice weekly from 3 days pre-surgery to 2 weeks post-surgery; once weekly from 3 to 8 weeks post-surgery; once every 2-4 weeks from 3 to 6 months post-surgery.
15. MRI: Includes T1-weighted imaging (T1W), T2-weighted imaging (T2W), Fluid-Attenuated Inversion Recovery (FLAIR), Diffusion Tensor Imaging (DTI), Diffusion-Weighted Imaging (DWI), and Magnetic Resonance Spectroscopy (MRS). Contrast-enhanced MRI may be used for pre-surgical planning to avoid blood vessels.
16. Early Withdrawal Visit: Subjects who discontinue from the study prematurely for any reason after completing cell therapy, or if the study is terminated prematurely, must complete an early withdrawal visit.

### 2 List of abbreviations and definition of terms

| Abbreviation | Description |
|---|---|
| AE | Adverse Event |
| ALB | Albumin |
| ALP | Alkaline Phosphatase |
| ALT | Alanine Aminotransferase |
| APTT | Activated Partial Thromboplastin Time |
| ASMs | Anti-Seizure Medications |
| AST | Aspartate Aminotransferase |
| BA | Basophils |
| BAI | Beck Anxiety Inventory |
| BNT-60 | Boston Naming Test |
| BVMT-R | Brief Visuospatial Memory Test-Revised |
| Ca | Calcium |
| CGE | Caudal Ganglionic Eminence |
| CI | Confidence Interval |
| Cl | Chloride |
| Cr | Creatinine |
| CRF | Case Report Form |
| CRO | Contract Research Organization |
| CT | Computed Tomography |
| DBIL | Direct Bilirubin |
| DSMB | Data and Safety Monitoring Board |
| DNA | Deoxyribonucleic Acid |
| DSA | Donor Specific Antibody |
| ECG | Electrocardiogram |
| eCRF | Electronic Case Report Form |
| EDC | Electronic Data Capture |
| EEG | Electroencephalogram |
| EP | Epilepsy |

| Abbreviation | Description |
|---|---|
| EO | Eosinophils |
| FDG-PET | Fluorodeoxyglucose-Positron Emission Tomography |
| FAS | Full Analysis Set |
| GABA | Gamma-Aminobutyric Acid |
| GLB | Globulin |
| GvHD | Graft-versus-Host Disease |
| HAMD | Hamilton Depression Rating Scale |
| Hb | Hemoglobin |
| HBV | Hepatitis B Virus |
| HCT | Hematocrit |
| HCV | Hepatitis C Virus |
| HGB | Hemoglobin |
| HIV | Human Immunodeficiency Virus |
| HR | Hazard Ratio |
| HTLV | Human T-lymphotropic Virus |
| ICF | Informed Consent Form |
| IEDs | Interictal Epileptiform Discharges |
| IL | Interleukin |
| ILAE | International League Against Epilepsy |
| INR | International Normalized Ratio |
| iPSC | Induced Pluripotent Stem Cell |
| K | Potassium |
| LITT | Laser Interstitial Thermal Therapy |
| LVFF | Left Ventricular Ejection Fraction (LVEF) |
| MGE | Medial Ganglionic Eminence |
| MGE-pINs | MGE-derived pallial GABAergic interneurons |
| MHC | Major Histocompatibility Complex |
| MO | Monocytes |
| MoCA | Montreal Cognitive Assessment |
| MRI | Magnetic Resonance Imaging |

| Abbreviation | Description |
|---|---|
| MRS | Magnetic Resonance Spectroscopy |
| MTLE | Mesial Temporal Lobe Epilepsy |
| Na | Sodium |
| NE | Neutrophils |
| nNOS | Neuronal Nitric Oxide Synthase |
| P | Phosphorus |
| PB | Peripheral Blood |
| pINs | Pallial GABAergic interneurons |
| PK | Pharmacokinetics |
| PLT | Platelets |
| PT | Prothrombin Time |
| PRA | Panel Reactive Antibody |
| PV | Parvalbumin |
| QOLIE-31 | Quality of Life in Epilepsy Inventory-31 |
| QTc | Corrected QT Interval |
| RAVLT | Rey Auditory Verbal Learning Test |
| RBC | Red Blood Cell Count |
| SAE | Serious Adverse Event |
| SOC | System Organ Class |
| SRC | Safety Review Committee |
| SS | Safety Set |
| SST | Somatostatin |
| TBIL | Total Bilirubin |
| TLR | Temporal Lobe Resection |
| TRAE | Treatment-Related Adverse Event |
| TPROT | Total Protein |
| UA | Uric Acid |
| UCB | Umbilical Cord Blood |
| ULN | Upper Limit of Normal |
| WBC | White Blood Cell Count |

| Abbreviation | Description |
|---|---|
| WHO | World Health Organization |
| γ-GT | Gamma-Glutamyl Transferase |

### 3 Introduction

#### 3.1 Background

Epilepsy (EP) is a chronic neurological disorder characterized by an imbalance between excitatory and inhibitory neuronal activity in the brain, leading to abnormal synchronous discharges within neural networks.[1,2] The clinical manifestations of these abnormal discharges are highly heterogeneous and depend on several key factors: the site of discharge onset, propagation patterns, brain maturity, comorbidities, sleep-wake cycles, and pharmacological interventions. These manifestations can affect multiple neurological domains, including sensory, motor, autonomic function, consciousness, emotion, memory, cognition, and behavior.[3] Epilepsy has become a significant global public health issue. Data from 2021 indicate that approximately 51.7 million people worldwide are affected by epilepsy.[4] The disease not only poses a direct threat to patient safety but also severely impacts their quality of life, social functioning, and economic status.

Based on the origin of seizures, focal epilepsy is the most common type, accounting for approximately 61% of all cases.[5–7] Its primary characteristic is that abnormal discharges originate in a specific brain region (epileptogenic zone) and subsequently spread to surrounding tissues.[8,9] In focal epilepsy, 60%-70% of seizures originate in the temporal lobe, with Mesial Temporal Lobe Epilepsy (MTLE) being the most prevalent.[6]

Anti-seizure medications (ASMs) play a crucial role in the treatment of epilepsy. Although three generations of ASMs have been successively applied in clinical practice with extensive accumulated experience, 20%-30% of epilepsy patients still fail to achieve effective seizure control through medication, eventually developing drug-resistant epilepsy.[10] This phenomenon is particularly common in MTLE, where approximately 70% of cases exhibit drug-resistant characteristics.[11]

Surgical resection or laser ablation are effective treatment modalities for drug-resistant MTLE.[12–14] However, such procedures may cause serious adverse effects, such as memory impairment, language deficits, and cognitive decline; therefore, they are rarely used for patients with bilateral MTLE.[15] In contrast, cell therapy offers a potential new therapeutic avenue that is safer and has fewer side effects for patients with drug-resistant MTLE, particularly those who are ineligible for surgical treatment.

#### 3.2 Study Rationale

The core pathophysiological features of EP include an excitatory-inhibitory imbalance, wherein the dysfunction of gamma-aminobutyric acid (GABA) receptors and their associated pathways plays a critical role in the pathophysiology of epilepsy. As the primary inhibitory neurotransmitter in the cerebral cortex, GABA maintains an inhibitory tone that balances neuronal excitation; disruption of this balance may trigger seizures. Consequently, the administration of GABA receptor antagonists can induce seizures, while drugs that enhance GABAergic transmission are utilized for anti-epileptic therapy.[2]

Pallial GABAergic interneurons (pINs) constitute a key cell population mediating GABA release within local neural circuits.[16] Among them, pINs derived from the medial ganglionic eminence

(MGE-pINs) play an essential regulatory role in cortical circuits, and their dysfunction is closely associated with neurological disorders.[17] The vast majority of pINs in the neocortex and hippocampus develop from the MGE and the caudal ganglionic eminence (CGE),[18] with the MGE primarily producing parvalbumin (PV)-positive and somatostatin (SST)-positive pINs. Interneuron abnormalities, particularly in those derived from the MGE, have been observed in various cases of epilepsy. Prolonged seizures can impair neuronal function, with GABAergic interneurons being particularly susceptible to damage; the loss of these interneurons leads to hyperexcitability of neuronal circuits during epileptogenesis. In animal models of epilepsy, SST- and PV-positive pINs are significantly reduced or dysfunctional,[19,20] and their selective activation can significantly attenuate seizures.[21,22] Similarly, a significant loss of MGE-pINs is observed in the hippocampal tissue of patients with MTLE.[23,24]

Stem cell therapy holds extraordinary potential for addressing unmet major needs in the treatment of human diseases. One particularly promising approach for epilepsy is enhancing inhibitory function in epileptic brain regions by transplanting new inhibitory neurons. Recent studies have shown that MGE-pINs derived from embryonic tissue, when transplanted into rodent epilepsy models, can migrate locally, survive long-term, generate mature pINs, and achieve functional integration with adult neural circuits,[25] thereby significantly suppressing seizures.[26–28] Notably, transplantation of CGE-pINs does not suppress seizures and may instead cause disinhibition effects.[29] Given that excitatory-inhibitory imbalance is central to the pathophysiology of epilepsy, and current surgical treatments (such as partial brain resection, laser ablation, and neurostimulation) primarily target epileptic networks on a macroscopic scale without directly correcting circuit-level abnormalities driving seizure generation, GABAergic interneuron transplantation emerges as a novel therapeutic strategy with the potential to rectify circuit abnormalities and restore normal regulatory function to the nervous system.

Preclinical studies have confirmed the feasibility of this therapeutic strategy. In a rodent model of MTLE established by intra-hippocampal injection of kainic acid (KA), the transplantation of human embryonic stem cell-derived MGE-pINs (hESC-MGE-pINs) sustainably suppressed mesial temporal lobe seizures. The transplanted interneurons were able to disperse locally, functionally integrate, survive long-term, and significantly reduce granule cell dispersion (a pathological hallmark of MTLE).[30]

A Phase I/II clinical trial (NCT05135091) targeted patients with drug-resistant unilateral mesial temporal lobe epilepsy using allogeneic hESC-MGE-pINs (NRTX-1001) for unilateral hippocampal transplantation. Immunosuppressants were administered starting one week prior to transplantation and continued for one year. Two-year follow-up data post-transplantation showed: in the low-dose group, the median reduction in seizures was 92% between 7-12 months post-treatment, with 80% of subjects achieving a >75% reduction; in the high-dose group, the median reduction was 78% between 4-6 months post-treatment, with 60% of subjects achieving a >75% reduction. Regarding safety, there were no serious complications, and mild immunosuppressive reactions were reversible; memory and quality of life remained stable or improved, and no abnormalities were seen on imaging. Another Phase I/II study (NCT06422923) also utilizes NRTX-1001 for bilateral transplantation in patients with bilateral mesial temporal lobe epilepsy.

##### 3.3 Benefit-Risk Assessment

Currently,ASMs and surgical resection are the primary treatment modalities for MTLE. However, ASMs show suboptimal efficacy in approximately 20%-30% of patients,[12] and may carry long-term side effects. While surgical intervention can be effective, it entails significant risks of serious neurological adverse reactions, such as memory impairment and cognitive decline.[17] Therefore, there is an urgent need to develop novel therapies that are safer and have fewer side effects.

Given that the core pathophysiological mechanism of MTLE involves an excitatory-inhibitory imbalance caused by the loss of inhibitory neurons,[1,2] supplementing specific GABAergic interneurons via stem cell transplantation would be of great help in fundamentally correcting this circuit abnormality. In relevant preclinical studies, the transplantation of hESC-MGE-pINs significantly suppressed seizures.[30] Furthermore, early clinical trials of similar products (NCT05135091) have demonstrated an acceptable safety profile and significant efficacy, with no serious adverse reactions observed.

Regarding ALC05 (allogeneic iPSC-derived GABAergic interneurons) developed in this study, no specific safety issues such as tumorigenicity were observed in related toxicological and pharmacodynamic studies, which also demonstrated good differentiation and integration capabilities. This cell therapy holds the promise of overcoming the limitations of existing treatments, offering patients a new avenue to restore normal regulatory functions of the nervous system. Through a rigorous phased design, a comprehensive risk monitoring plan, and long-term follow-up arrangements, this study maximally mitigates potential risks associated with surgery and cell transplantation. Therefore, for patients with drug-resistant MTLE, the expected benefits of intracranial transplantation of ALC05 are determined to outweigh the risks.

### 4 Study Objectives and Endpoints

##### 4.1 Study Objectives

To explore the safety and preliminary efficacy of a single intracranial injection of ALC05 in patients with unilateral MTLE, and to evaluate the therapeutic effects of different doses of ALC05 injection.

##### 4.2 Study Endpoints

###### 4.2.1 Primary Endpoint

The frequency and severity of adverse events (AEs) and serious adverse events (SAEs) within 12 months post-surgery.

###### 4.2.2 Secondary Endpoints

**Efficacy:**

1. Cell Engraftment and Survival: Changes in metabolic and functional status of brain tissue assessed by Fluorodeoxyglucose-Positron Emission Tomography (FDG-PET) and Magnetic Resonance Spectroscopy (MRS) compared to baseline (pre-transplantation).
2. Responder Rate: The percentage of subjects with a ⩾50% reduction in seizure frequency compared to baseline.
3. Seizure Frequency: Change in seizure frequency (number of seizures per 28 days) compared to baseline, based on seizure diaries and EEG; including the frequency of all seizure events and disabling seizures.

##### 4.2.3 Exploratory Endpoints

1. Quality of Life Assessment: Change in Quality of Life in Epilepsy Inventory-31 (QOLIE-31) scores compared to baseline.
2. Neurocognitive Function Assessment: Changes in scores (or results) compared to baseline for the Montreal Cognitive Assessment (MoCA), Rey Auditory Verbal Learning Test (RAVLT), Boston Naming Test (BNT-60), and Brief Visuospatial Memory Test-Revised (BVMT-R).
3. Mood Disorder Assessment: Changes in Hamilton Depression Rating Scale (HAMD) and Beck Anxiety Inventory (BAI) scores compared to baseline.

### 5 Study Population

#### 5.1 Sample Size

This study plans to enroll 12 patients with MTLE who meet the inclusion criteria and do not meet the exclusion criteria. All enrolled subjects will be randomly divided into either low dose or high dose group at a 1:1 ratio. To minimize risks to subjects at each dose level, the first subject in each dose group will be monitored for safety for at least 3 months post ALC05 injection and show acceptable safety and tolerability, before the remaining patients can be enrolled. The safety data will be reviewed by the Independent Data Safety Monitoring Board (DSMB) (defined as no occurrence of serious adverse events or adverse events leading to treatment discontinuation; and no clinically significant abnormal changes in safety indicators such as laboratory tests and vital signs).

#### 5.2 Inclusion Criteria

Patients are eligible for inclusion in the study only if they meet all of the following criteria:

1. Capable of signing the informed consent form and complying with the study protocol;
2. Male or female, aged 18 to 75 years (inclusive);
3. Long-term scalp electroencephalogram (EEG) monitoring, cranial imaging, and clinical symptoms indicate Mesial Temporal Lobe Epilepsy (MTLE);
4. Currently receiving adequate doses, sufficient duration, and rational use of ≥2 anti-seizure medications (ASMs) approved by the NMPA, with a treatment duration of ≥1 month, yet failing to achieve seizure control;

5. Average seizure frequency of ≥2 seizures per 28 days during the 6 months prior to the screening visit;
6. Investigator considers the patient suitable for Temporal Lobe Resection (TLR) or Laser Interstitial Thermal Therapy (LITT).

#### 5.3 Exclusion Criteria

Subjects will be excluded from the study if they meet any of the following criteria:

1. Patients with concomitant conditions as follows: a) Other significant brain structural abnormalities (e.g., brain tumors, vascular malformations, arachnoid cysts, tuberous sclerosis, cortical dysplasia, etc.); b) History of Status Epilepticus within 1 year prior to screening (based on ILAE criteria, Trinka 2015, determined by the Principal Investigator; history of seizure clusters is permitted); c) Autoimmune epilepsy, with relevant serum antibodies detected within 3 years prior to screening or at baseline (accepted methods include Labcorp 505490, Mayo Clinic EPS2, or similar tests approved by the Sponsor); d) Progressive neurological diseases (e.g., multiple sclerosis, primary mitochondrial diseases, other neurodegenerative diseases); e) History of severe cranial diseases (e.g., stroke, traumatic brain injury, subarachnoid hemorrhage); f) Severe cognitive impairment, intellectual disability, or psychiatric disorders (e.g., schizophrenia, severe depression) that prevent completion of study procedures or provision of valid informed consent; g) History of suicide attempts or suicidal ideation within 12 months prior to enrollment;
2. Unstable vital signs at screening and/or pre-surgery: a. Heart rate <50 bpm or >105 bpm; b. Systolic Blood Pressure (SBP) <90 mmHg or >160 mmHg; Diastolic Blood Pressure (DBP) <60 mmHg or >100 mmHg; c. Respiratory rate <12 breaths/min or >20 breaths/min; d. Resting pulse oxygen saturation (SpO2) <94%; e. Body temperature >100.4°F (38.0°C);
3. Estimated Glomerular Filtration Rate (eGFR) <30 ml/min/1.73m², and uncorrectable;
4. Abnormal liver function: ALT or AST >3 times the upper limit of normal (ULN), and uncorrectable;
5. Hematological abnormalities: e.g., Hematocrit <25%, White Blood Cell count <2,500/µl, or Platelet count <100,000/µl, and uncorrectable;
6. Abnormal coagulation function: INR >1.3, and uncorrectable;
7. Patients with autoimmune diseases;
8. Long-term use of immunosuppressants, such as systemic corticosteroids, TNFα inhibitors, etc.;
9. History of prior organ transplantation;
10. Patients with diabetes mellitus (due to higher risk of underlying disease exacerbation or new infections);
11. Positive for Human Immunodeficiency Virus (HIV), Hepatitis B Virus (HBV), or Hepatitis C Virus (HCV);
12. Positive Panel Reactive Antibody (PRA);
13. Patients who have received cell or gene therapy (autologous or allogeneic);
14. History of non-pharmacological treatments (e.g., surgical resection, thermal ablation, radiation therapy, VNS, DBS);
15. Participation in investigational treatment or device trials at the time of signing informed consent or within 30 days after obtaining informed consent;

16. Patients unable to undergo MRI and PET-CT examinations;
17. Allergy to radiocontrast agents and unable to be managed with premedication;
18. History of allergy to methotrexate, ruxolitinib, or penicillin, or non-sensitive to tacrolimus based on genetic testing;
19. Progressive malignancy within the past 5 years (excluding basal cell carcinoma);
20. Life expectancy <1 year;
21. Pregnant or breastfeeding women;
22. Women of Childbearing Potential (WOCBP) who cannot commit to strict adherence to effective contraceptive measures;
23. Any other patients deemed unsuitable for participation in this clinical study by the Investigator.

#### 5.4 Screen Failure

Screen failure is defined as a subject who has consented to participate in the clinical study but subsequently does not receive the investigational medicinal product. For screen-failed subjects, at a minimum, demographic data and the reason for screen failure must be collected.

#### 5.5 Subject Withdrawal Criteria

##### 5.5.1 Investigator-Initiated Withdrawal

This refers to instances where the Investigator determines that an enrolled subject is no longer suitable to continue in the study, and consequently, the Investigator decides to withdraw the subject:

- Poor subject compliance, which impacts the assessment of safety and tolerability;
- The subject experiences an adverse event which, upon analysis and management, makes it unsuitable for them to continue in the study;
- Other circumstances where the Investigator deems the subject unsuitable to continue in the study.

##### 5.5.2 Subject-Initiated Withdrawal

1. Subjects have the right to withdraw from the study at any time for any reason;
2. If a subject does not explicitly request to withdraw but fails to attend visits, receive the cell product, or cooperate with assessments as required by the study protocol, and the Investigator is unable to contact the subject on at least three occasions between the current and subsequent visit periods, the subject may be considered "lost to follow-up." This type of lost to follow-up is also classified as "withdrawal."

##### 5.5.3 Procedures for Subject Withdrawal

If a subject withdraws due to an adverse event, these subjects will be kept under close medical observation until the adverse event recovers, stabilizes, or reaches another definitive outcome. The detailed outcome status will be handled according to the procedures for "Adverse Event Follow-up." The specific reason for the subject's withdrawal must be determined as completely and accurately as possible. The Investigator is required to document the specific reason for subject withdrawal in both the source documents and the electronic Case Report Form (eCRF).

#### 5.6 Study Suspension/Discontinuation

##### 5.6.1 Principles for Suspension/Termination

1. During the study, the Investigators and the Sponsor (or Collaborator) may discuss and decide to terminate the study based on preliminary data. All decisions made following such discussions shall be clearly documented in the meeting minutes.

2. Serious safety issues occur during the trial, such as the occurrence of > 2 Serious Adverse Events (SAEs).

3. Major errors in the clinical study protocol are discovered during the study, making it difficult to evaluate the effects of the cell product; or major deviations occur during the implementation of a well-designed protocol, making it difficult to evaluate the effects of the cell product if continued.

4. Reasons related to the Sponsor/Collaborator, such as insufficient funding or patent disputes, or changes in national pharmaceutical development policies.

5. The drug regulatory authorities or the Ethics Committee (EC) require the suspension or termination of an approved study.

Once the issues causing the suspension or termination—such as cell product safety or protocol compliance—are resolved and approval is obtained from the Sponsor/Collaborator and the Ethics Committee, the study may resume. Any party deciding to suspend or terminate the study must immediately issue a written notice to the other parties (including but not limited to the Sponsor/Collaborator, Investigators, and Ethics Committee) and provide relevant reasons. Based on the evaluation results, the clinical study protocol may be amended to reduce safety risks to subjects. Amendments typically include revisions to inclusion criteria (e.g., excluding subject populations with a higher risk of specific adverse events), dose reduction, adjustments to product formulation or administration methods, or improvements to the subject safety monitoring plan. The study may be considered for resumption after the protocol has been adjusted and improved.

##### 5.6.2 Lost to Follow-up

A subject will be considered lost to follow-up if they repeatedly fail to return for scheduled visits or if the study site is unable to contact them.

If a subject fails to return to the study site for required study visits, the following measures must be taken:

- The site must attempt to contact the subject to reschedule the missed visit as soon as possible, counsel the subject on the importance of adhering to the visit schedule, and determine whether the subject wishes and/or should continue in the study;
- Before confirming a subject as lost to follow-up, the Investigator or designee must make every reasonable effort to contact the subject (where possible, making 3 telephone calls and, if necessary, sending a certified letter to the subject's last known mailing address or using equivalent methods). All attempts should be documented in the subject's medical records;
- If contact cannot be established after all attempts, the subject will be considered withdrawn from the study.

For subjects lost to follow-up, every effort should be made to contact them to collect information regarding any new treatments received after terminating this study.

### 6 Study Design

#### 6.1 Overall Design

This is a single-center, randomized, double-blind, investigator-initiated clinical study designed to evaluate the safety and preliminary efficacy of ALC05 injection. ALC05 is a gene-edited, low-immunogenicity iPSC-derived GABAergic interneurons.

This study plans to recruit 12 patients with MTLE who meet the inclusion criteria and do not meet the exclusion criteria. After adequate informed consent, subjects will sign the Informed Consent Form. All enrolled subjects will be randomly divided into either low dose or high dose group at a 1:1 ratio. To minimize risks to subjects at each dose level, the first subject in each dose group will be monitored for safety for at least 3 months post ALC05 injection and show acceptable safety and tolerability, before the remaining patients can be enrolled. The safety data will be reviewed by the Independent DSMB (defined as no occurrence of serious adverse events or adverse events leading to treatment discontinuation; and no clinically significant abnormal changes in safety indicators such as laboratory tests and vital signs). Subjects will be aged 18 to 75 years, with a confirmed diagnosis of MTLE at screening, and have failed to achieve seizure control despite adequate treatment with at least two ASMs at appropriate doses for a sufficient duration. The complete inclusion and exclusion criteria are detailed in the relevant section. Subjects will complete screening, baseline, and pre-operative visits before surgery, and receive a post-operative assessment on Day 7 post-transplantation. Seizure frequency and clinical rating scale assessments will be conducted via on-site visits during the study period. The total follow-up period post-transplantation is 1 year. Upon completion of their follow-up period in this study, subjects will enter a post-study long-term safety follow-up, continuing until the subject's death or loss to follow-up. After the end of treatment, subjects will be followed up once at years 1, 2, 3, 5, 8, and 10, via outpatient visits (follow-up indicators include imaging examinations, responder rate, seizure frequency, and seizure classification); thereafter, they will be followed up once every 5 years via telephone calls to ascertain their survival status. This long-term follow-up plan is not included within the 12-month visits of this study, and is for subject information only.

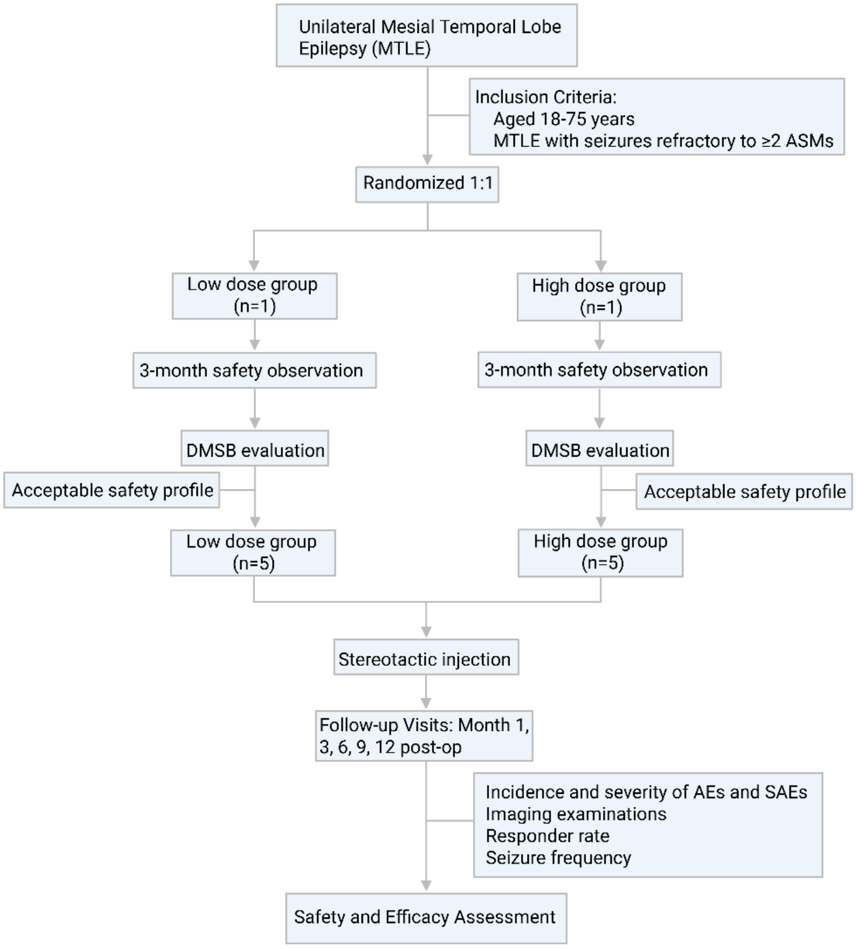

**Figure 1 Flow Diagram of the Trial**

#### 6.2 Randomization

##### 6.2.1 Randomization Strategy

Considering that this is a First-in-Human (FIH) trial, and for safety reasons, the first subject in each dose cohort will not be randomized but will be directly assigned to the corresponding dose group. Enrollment and randomization of subsequent subjects will commence only after the first subject in each dose cohort completes at least 3 months of safety observation and safety is confirmed as acceptable by an independent DSMB (defined as no occurrence of serious adverse events or adverse events leading to treatment discontinuation; and no clinically significant abnormal changes in safety indicators such as laboratory tests and vital signs).

Following the completion of the preliminary safety assessment for the first subject in each dose cohort, the subsequent 10 subjects will be randomized to the two dose groups using a simple randomization method, aiming for an allocation ratio of approximately 1:1. Considering the small sample size (N=10) and exploratory nature of this study, no stratification factors will be set. The randomization list will be generated and maintained by a statistician independent of the study team and will be used to guide subject dose assignment.

##### 6.2.2 Randomization Process

Randomization for the subsequent 10 subjects will be automatically implemented via the randomization module of the Viedoc Electronic Data Capture (EDC) system. This module features encrypted storage, access control, and comprehensive audit trail functions to ensure the accuracy of the randomization process and the maintenance of blinding.

The randomization sequence will be generated by a biostatistician independent of the study team using SAS Version 9.4 or other validated statistical software. The sequence will employ simple randomization with a 1:1 allocation ratio to the low-dose and high-dose groups. The generated randomization sequence will be encrypted and uploaded to the Viedoc EDC system's randomization module, where the algorithm and sequence will be securely stored.

After the Investigator completes the subject's screening assessment and enters all baseline data in the Viedoc EDC system, the system will automatically verify compliance with the preset inclusion/exclusion criteria. Upon confirmation that the first subject has completed the preliminary safety assessment and the results are acceptable, the Investigator may click the "Randomize" button in the system. The Viedoc system will automatically assign the treatment group based on the preset randomization sequence and generate a unique randomization number for the subject. The system will simultaneously display a corresponding drug package number for the subject, which the Investigator will use to dispense the study drug.

The Investigator will dispense the corresponding study drug from the investigational pharmacy according to the drug package number displayed by the Viedoc system. The drug package label will correspond one-to-one with the system-assigned randomization number and will only display the subject number and usage instructions, without revealing specific dose information, thereby maintaining blinding. The pharmacist must record drug dispensation information in the system, including drug number, dispensation date, and quantity.

Treatment assignment information will be securely stored and encrypted within the Viedoc system. Clinical Investigators will have system access permissions to view treatment assignments for correct drug administration and safety management; however, all blinded assessors (including seizure frequency assessors, scale assessors, and imaging analysts) will have system access permissions set to prevent viewing treatment assignment information and drug package numbers, to strictly protect blinding. An emergency unblinding function will be configured within the system, recording the time, reason, and operator of all unblinding actions. The system will automatically lock randomized subjects to prevent duplicate operations.

All randomization operations (including system verification, assignment results, drug dispensation) will be automatically recorded in the Viedoc system's audit trail, including operator, time of operation, assignment results, etc. The Sponsor and Investigators will regularly review randomization logs, and monitors will verify the consistency of randomization records with actual drug dispensation during monitoring visits to ensure the completeness and accuracy of the randomization process.

#### 6.3 Blinding

##### 6.3.1 Blinding Design

This study employs a modified, prospective, randomized, double-blind design. Specifically, the study blinds subjects and investigators, while also maintaining blinding for endpoint assessors.

This design aims to achieve double-blinding for investigators and subjects by having a central pharmacy uniformly prepare cell suspensions identical in appearance and volume, thereby maximizing the elimination of subjective bias in efficacy assessment while ensuring operational safety. Key efficacy measures, such as seizure diaries and quality of life scales for epilepsy (e.g., QOLIE-31), are susceptible to subjective expectations. Concealing specific dose assignments (low dose or high dose) from subjects and clinical investigators effectively eliminates subjects' psychological expectations and reporting bias, and prevents unconscious guidance from investigators that could lead to assessment bias, thereby ensuring the objectivity and authenticity of efficacy data.

Since the central pharmacy is responsible for preparing cell suspensions of different doses with identical appearance, volume, packaging, and external labels according to the randomization scheme, clinical investigators only receive and use visually identical drugs for surgery and administration. Therefore, the clinical investigator team directly responsible for subject surgery, administration, and safety management (including PI and Sub-I) does not need to know the specific treatment assignment information, thus achieving blinding for clinical investigators for the first time in this study. This fundamentally cuts off potential pathways for treatment information leakage to the efficacy assessment process. All personnel responsible for assessing and adjudicating primary and secondary efficacy endpoints (including seizure frequency assessors, scale scorers, and imaging analysts) must remain completely blinded to treatment assignments. This measure maximally reduces assessor bias, ensuring the impartiality and scientific validity of efficacy determination.

For the first subjects in each dose group, who are directly assigned without randomization, the study will still implement subject blinding and endpoint assessor blinding strategies to ensure all subjects follow unified assessment standards throughout the study period.

##### 6.3.2 Management of Blinded Personnel

The following personnel will remain blinded to the specific dose assignment information throughout the trial: all subjects (including the first non-randomized subjects); PI; Sub-I; efficacy assessment personnel (seizure frequency assessment physician; scale assessors; imaging assessment physician); statistical analysis personnel.

The following personnel, due to their job responsibilities (administration, surgery, or safety monitoring), will be aware of the subject's treatment dose assignment information: central pharmacy pharmacist (responsible for receiving randomization information and preparing visually identical drugs); Sponsor Clinical Monitor; Sponsor Medical Monitor; Sponsor-designated unblinded medical safety monitor (not involved in routine clinical care or efficacy assessment, responsible for emergency unblinding for serious adverse events and overall safety review).

##### 6.3.3 Blinding Management Procedures

The following measures will be taken during the study to maintain the blinding of subjects and endpoint assessors, ensuring the objectivity and reliability of study results.

Cell products for different dose groups will be prepared by the central pharmacy according to the randomization scheme. The cell suspensions dispensed to clinical investigators will be identical in dosage form, appearance, color, volume, containment vessel, and external packaging label. Drug package labels will only display necessary information such as the study protocol number, subject randomization number, unified usage and dosage instructions, storage conditions, and expiration date, strictly prohibiting the display of specific dose information or any hint of treatment assignment.

Central pharmacy pharmacists must strictly avoid revealing dose information in communications with Investigators, and drug preparation should be conducted in a separate room, ensuring that blinded personnel are not present. During the informed consent process, subjects in each dose group will only be informed that they will receive investigational drug treatment, and subsequent subjects will be informed that they will be randomized to one of two dose groups, but neither will be told the specific dose or assignment result. All subjects will be emphasized not to attempt to guess or inquire about their treatment assignment throughout the study period, and will be reminded not to disclose any treatment-related information during efficacy assessments. If a subject accidentally discloses such information, the assessor should immediately record and report it to the Principal Investigator. Imaging data will be de-identified by specific personnel, removing personal identity information and treatment assignment information, only retaining subject randomization number and examination time points. Imaging experts will perform blinded readings according to unified standards, with critical imaging data double-read by two independent experts. In cases of disagreement, a third senior expert will arbitrate to ensure the accuracy and consistency of assessment results.

The Sponsor's Clinical Monitor will verify the maintenance of blinding during each monitoring visit, including checking if the central pharmacy's drug preparation and dispensation records are compliant, whether any blinding breaches occurred, checking the effectiveness of physical separation, and spot-checking assessors' knowledge of treatment assignment (inquiring if subjects accidentally disclosed information, checking if materials hinting at assignments were inadvertently placed in assessment rooms).

The Sponsor's Medical Monitor and Sponsor-designated unblinded medical safety monitor will only access treatment assignment information when necessary (e.g., for emergency unblinding due to a serious adverse event), and must strictly adhere to information isolation procedures to ensure that the information they know is not disclosed to any blinded personnel (including clinical investigators, endpoint assessors, and statisticians).

The Viedoc EDC system's data administrator will regularly check if user access permissions are correctly set, confirming that blinded personnel accounts cannot access modules containing treatment assignment information. If the system indicates any access permission anomalies or unauthorized accounts attempting to access treatment assignment information, it must be promptly investigated and reported.

###### 6.3.4 Unblinding Procedures

All blinded group assignment information will only be unblinded after database lock and completion of statistical analysis.

Emergency unblinding is limited to extremely special circumstances that threaten the subject's life safety or may affect the subject's critical clinical treatment. It must follow pre-approved emergency unblinding procedures and be executed with the approval of the Sponsor's Medical Monitor.

### 7 Study Intervention

#### 7.1 Administration Regimen

##### 7.1.1 Cell Product Information

| Item | Description |
|---|---|
| **Investigational Treatment Name** | ALC05 Cell Injection |
| **Type** | Cell therapy and cell-based products |
| **Dosage Form** | Cell injection (suspension for injection) |
| **Strength/Specification** | $1 \times 10^7$ cells/mL, 1 mL/vial |
| **Storage Conditions** | -196°C to -130°C |

| | |
|---|---|
| **Administration Site** | Hippocampus |
| **Route of Administration** | Stereotactic injection into the hippocampus |
| **Packaging and Labeling** | Each package will be labeled in accordance with local regulatory requirements. |

##### 7.1.2 Receipt, Dispensing, and Storage of Cell Product

Only subjects enrolled in the study are eligible to receive surgical transplantation treatment with ALC05 Cell Injection, and only authorized study site personnel may dispense or administer the ALC05 Cell Injection. All investigational products or related items must be stored in a secure, environmentally controlled, and monitored (manually or automatically) area according to the storage conditions indicated on the label. Access to the investigational treatment product must be restricted to the Investigator and authorized study site personnel.

The Investigator, institution, or head of the medical facility (as applicable) is responsible for the accountability, reconciliation, and record maintenance (i.e., records of receipt, reconciliation, and final disposition) of the investigational treatment.

Further guidance and information regarding specific drug preparation, administration procedures, and the final disposition of unused investigational medicinal products can be found in the Pharmacy Manual.

##### 7.1.3 Preparation of Cell Product

All cell products are manufactured at a GMP-compliant facility and undergo rigorous quality control testing and release testing. Cell batches that pass release testing will be cryopreserved and subsequently prepared for transport according to simulated and validated packaging and shipping procedures. The cell product will be delivered directly to the operating room via a cold chain transport system under temperature-controlled conditions throughout the entire journey, with continuous temperature monitoring implemented during transport.

On the day of surgery, the ALC05 cell product will undergo thawing and preparation procedures under strict aseptic conditions to ensure compliance with clinical standards for transcranial stereotactic injection. The entire transport and handling process strictly adheres to full traceability management to ensure that the identity, biological activity, and quality integrity of the cell product are fully guaranteed. The usage of all ALC05 injections must be accurately documented.

##### 7.1.4 Method of Administration

On the day of transplantation treatment, the following steps must be completed for the confirmed subject:

i. On the day of surgery (D1), the subject will undergo general anesthesia, and a Mayfield head clamp will be fixed to the subject's skull.

ii. Pre-operative MRI/CT data of the subject will be imported into the Huake Robot Workstation for image fusion and registration.

iii. Upon successful registration, the pre-planned injection trajectory will be retrieved. The robotic arm will automatically maneuver and position itself to the planned entry point and angle.

iv. Along the locked path of the robotic arm, the guide cannula will be inserted to the predetermined depth, followed by the removal of the stylet from within the cannula.

v. After measuring the length of the cell injection needle, the depth stop (limiter) will be installed and adjusted. The injection needle will then be inserted through the guide cannula to the target site.

vi. A micro-pump will be used to assist in pushing the syringe at a constant speed, while simultaneously retracting the injection system along the trajectory.

Post-operatively, the subject will undergo rigorous perioperative monitoring and immunosuppressive therapy. Comprehensive contingency plans will be in place. All examinations will be completed according to the Study Schedule, and changes in the patient's vital signs and neurological signs will be recorded. On Day 2 (D2), a CT/MRI scan will be performed to assess the transplantation site for the presence of hematoma.

#### 7.2 Concomitant Medications/Treatments

All medications or vaccines used (including over-the-counter drugs or prescription drugs, vitamins and/or herbal supplements) and all procedures performed by the subject from prior to the cell transplantation surgery (endeavor to collect 1-3 months of medication/treatment information pre-surgery) until the End of Treatment visit should be recorded in the eCRF. In addition, previous treatments for underlying diseases should be recorded. If a subject requires concomitant use of other medications during the study for therapeutic reasons, it must be approved by the Investigator, and administered under the Investigator's guidance.

All concomitant medications and concomitant treatments must be recorded in the concomitant medication section of the eCRF.

The record should include the following information:

- Name;
- Reason for use;
- Start and stop dates;

- Dose information, including dosage and frequency.

##### 7.2.1 Immunosuppressive Regimen

Immunosuppressive Regimen in This Study is as Follows:

1. Tacrolimus Administration: Oral tacrolimus will be administered from 3 days pre-surgery to 8 weeks post-surgery, with an initial dose of 0.075 mg/kg/day, twice daily (bid). The dose will be adjusted based on the patient's blood trough concentration, aiming for a target concentration of 5-10 ng/mL. Blood levels will be monitored twice weekly in the early postoperative period, followed by regular monitoring until the end of immunosuppressive drug therapy.

2. Tacrolimus Tapering: Doses will be gradually tapered starting from 3 months post-surgery. If imaging and clinical indicators meet the specified criteria, tacrolimus will be discontinued at 6 months post-surgery.

3. Prophylaxis for Immunosuppressive Side Effects:

    a. Pneumocystis Pneumonia (PCP) Prophylaxis: Co-trimoxazole (trimethoprim/sulfamethoxazole), standard dose once daily, or double dose three times weekly, continued for 6 months.

    b. Herpes Simplex Virus (HSV)/Varicella-Zoster Virus (VZV) Prophylaxis: Aciclovir 400 mg, twice daily, continued for 3 months.

    c. Cytomegalovirus (CMV) Prophylaxis: For donor positive/recipient negative pairs, valganciclovir will be administered for 3 months or as per the study site's protocol.

    d. Fungal Prophylaxis: Fluconazole or an equivalent drug will be administered for 3 months or as per the study site's protocol.

    e. Gastrointestinal Protection: Proton pump inhibitors (PPIs) will be used in the early phase of calcineurin inhibitor therapy.

4. Treatment of Rejection/Neuroinflammatory Reaction:

    a. Exclusion of infection (via MRI, and lumbar puncture if necessary).

    b. Methylprednisolone 500–1000 mg IV, once daily, for 3 consecutive days, followed by gradual tapering.

    c. Adjust tacrolimus dose to the upper limit of the target range.

    d. Add mycophenolate mofetil capsules 0.5 g bid until the end of treatment.

##### 7.2.2 Prohibited and Restricted Medications

In general, any concomitant medication/treatment required by the subject is permitted, with the exception of the following:

- Prohibited Epilepsy Treatments During the Study: Subjects are prohibited from receiving the following treatments during the study period, including but not limited to TLR, LITT, other autologous/allogeneic cell transplantation/infusion, or participation in other clinical studies indicated for epilepsy.

- Prohibition on Other Studies: Participation in other clinical studies involving cells, drugs, or devices is prohibited for 2 years post-surgery.

##### 7.2.3 Permitted Concomitant Medications

For medications taken prior to signing the Informed Consent Form (ICF), the current regimen may be maintained. For new symptoms or conditions emerging during the study, medications may be administered based on the judgment of the Investigator or designee, including but not limited to the following situations:

- Use of ASMs: The use, dosage, and administration of other ASMs will be determined by the Investigator based on the subject's condition.

- Supportive Therapy: The Investigator is permitted to use supportive therapies in accordance with the clinical site's medical guidelines. These include, but are not limited to, antibiotics, platelet transfusions, plasma transfusions, bronchodilators, epinephrine, antihistamines, intracranial decompression drugs, or glucocorticoids.

#### 7.3 Treatment Compliance

During the recruitment and screening phase, the Investigator will inform subjects about the study's purpose, basic information about the investigational drug, study protocol, study procedures, dosing regimen (e.g., dose, administration method, cycle), clinical observations, frequency and procedures for biological sample collection, potential risks of participation, compensation, and indemnity. This is to ensure subjects are fully informed and voluntarily participate, thereby enhancing subject compliance. During the informed consent discussion, the Investigator must emphasize compliance to subjects, explaining that compliance is essential for the subject's safety and the study's efficacy. The Investigator will guide subjects on attending planned study visits to reinforce compliance. Subjects should be instructed to contact the Investigator if they cannot follow the protocol's planned follow-up for any reason. During the study, if a subject shows poor compliance, the Investigator should identify the reason, actively take appropriate measures (e.g., emphasizing the importance of protocol compliance to the subject), and fully document any non-compliance, its reasons, and the corresponding measures taken.

##### 7.4 Post-Study Treatment

After the completion of the study, the investigational product will generally no longer be provided to subjects.

### 8 Study Procedures and Assessments

#### 8.1 Visit Schedule

The Investigator should instruct subjects to attend outpatient or inpatient follow-up visits according to the schedule specified in the Study Schedule.

##### 8.1.1 Informed Consent

The Investigator shall explain the basic content of the study to each subject, including the study objectives, basic information about the cell product, study procedures, administration methods, potential risks of participation, compensation, and any other information the subject wishes to know. After full consideration and confirmation that the subject fully understands the study process and agrees to participate, they shall sign the written ICF. All subjects will be provided with a copy of their signed and dated ICF.

##### 8.1.2 Visit 1 and Visit 2: V1-V2 (Screening/Baseline Period D-30~D-1)

Subjects may participate in screening after signing the ICF. For procedures and assessments during Visit 1 (Screening Period), please refer to the Study Flowchart. The Investigator will screen subjects based on all data obtained during Visit 1. If a subject meets the inclusion criteria and does not meet the exclusion criteria, the subject will proceed to the next visit. The Investigator should educate subjects entering the next visit about adhering to the study protocol. If a subject does not meet the relevant screening requirements, they will be considered a screen failure, and the reason for screen failure must be recorded.

All screening/baseline procedures and assessments are detailed in the Study Schedule. After signing the ICF, subjects must complete all required examinations during the screening/baseline period. If they meet all inclusion criteria and do not meet any exclusion criteria, they are successfully enrolled in the study.

The following examinations must be completed, with content as described below for each item:

- Demographics, Medical/Treatment History, Physical Examination, Vital Signs
- PRA, DSA, Serum Pregnancy Test (WOCBP only), 12-Lead ECG, Hematology, Blood Biochemistry, CRP, Coagulation Panel, Urinalysis, Stool Routine + Occult Blood, EEG, Cytokines, Immune Panel, Peripheral Blood T-cell Subsets, Tumor Markers
- MoCA, RAVLT, QOLIE-31, BNT-60, BVMT-R, HAMD, BAI

- Brain CT Scan (General), Brain MRI, Brain PET-CT
- Adverse Events, Concomitant Medications/Treatments

##### 8.1.3 Visit 3: V3 (Day of Surgery D1)

Following completion of pre-operative preparations, intracranial stereotactic injection surgery will be performed, and the following examinations will be completed:

- Vital Signs, 12-Lead ECG
- Brain CT Scan (General), Brain MRI
- Adverse Events, Concomitant Medications/Treatments
- The Investigator may add the following examinations based on the subject's clinical presentation: Serum Pregnancy Test (WOCBP only), Physical Examination, Hematology, Blood Biochemistry, CRP, Coagulation Panel, Urinalysis, Stool Routine + Occult Blood, EEG, PRA, DSA, Cytokines, Immune Panel, Peripheral Blood T-cell Subsets, Tumor Markers

##### 8.1.4 Visit 4: V4 (D2)

The following examinations must be completed, with content as described below for each item:

- Vital Signs, 12-Lead ECG
- Brain MRI/CT
- Adverse Events, Concomitant Medications/Treatments
- The Investigator may add the following examinations based on the subject's clinical presentation: Serum Pregnancy Test (WOCBP only), Physical Examination, Hematology, Blood Biochemistry, CRP, Coagulation Panel, Urinalysis, Stool Routine + Occult Blood, EEG, PRA, DSA, Cytokines, Immune Panel, Peripheral Blood T-cell Subsets, Tumor Markers

##### 8.1.5 Visit 5: V5 (D8 Discharge, Window: ±3 days)

The following examinations must be completed, with content as described below for each item:

- Vital Signs, 12-Lead ECG
- Adverse Events, Concomitant Medications/Treatments
- The Investigator may add the following examinations based on the subject's clinical presentation: Serum Pregnancy Test (WOCBP only), Physical Examination, Hematology,

Blood Biochemistry, CRP, Coagulation Panel, Urinalysis, Stool Routine + Occult Blood, EEG, PRA, DSA, Cytokines, Immune Panel, Peripheral Blood T-cell Subsets, Tumor Markers

##### 8.1.6 Visit 6: V6 (Post-op M1, Window: ±7 days)

The following examinations must be completed, with content as described below for each item:

- Vital Signs
- Seizure Frequency and Type
- MoCA, RAVLT, QOLIE-31, BNT-60, BVMT-R, HAMD, BAI
- Adverse Events, Concomitant Medications/Treatments
- The Investigator may add the following examinations based on the subject's clinical presentation: Serum Pregnancy Test (WOCBP only), Physical Examination, Hematology, Blood Biochemistry, CRP, Coagulation Panel, Urinalysis, Stool Routine + Occult Blood, EEG, PRA, DSA, Cytokines, Immune Panel, Peripheral Blood T-cell Subsets, Tumor Markers

##### 8.1.7 Visit 7: V7 (Post-op M3, Window: ±7 days)

The following examinations must be completed, with content as described below for each item:

- Vital Signs
- Seizure Frequency and Type
- MoCA, RAVLT, QOLIE-31, BNT-60, BVMT-R, HAMD, BAI
- Adverse Events, Concomitant Medications/Treatments
- The Investigator may add the following examinations based on the subject's clinical presentation: Serum Pregnancy Test (WOCBP only), Physical Examination, Hematology, Blood Biochemistry, CRP, Coagulation Panel, Urinalysis, Stool Routine + Occult Blood, EEG, PRA, DSA, Cytokines, Immune Panel, Peripheral Blood T-cell Subsets, Tumor Markers

##### 8.1.8 Visit 8: V8 (Post-op M6, Window: ±7 days)

The following examinations must be completed, with content as described below for each item:

- Vital Signs
- Seizure Frequency and Type

- MoCA, RAVLT, QOLIE-31, BNT-60, BVMT-R, HAMD, BAI
- Adverse Events, Concomitant Medications/Treatments
- The Investigator may add the following examinations based on the subject's clinical presentation: Serum Pregnancy Test (WOCBP only), Physical Examination, Hematology, Blood Biochemistry, CRP, Coagulation Panel, Urinalysis, Stool Routine + Occult Blood, EEG, PRA, DSA, Cytokines, Immune Panel, Peripheral Blood T-cell Subsets, Tumor Markers

###### 8.1.9 Visit 9: V9 (Post-op M9, Window: ±7 days)

The following examinations must be completed, with content as described below for each item:

- Vital Signs
- Seizure Frequency and Type
- MoCA, RAVLT, QOLIE-31, BNT-60, BVMT-R, HAMD, BAI
- Adverse Events, Concomitant Medications/Treatments
- The Investigator may add the following examinations based on the subject's clinical presentation: Serum Pregnancy Test (WOCBP only), Physical Examination, Hematology, Blood Biochemistry, CRP, Coagulation Panel, Urinalysis, Stool Routine + Occult Blood, EEG, PRA, DSA, Cytokines, Immune Panel, Peripheral Blood T-cell Subsets, Tumor Markers

###### 8.1.10 Visit 10: V10 (Post-op M12, Window: ±7 days)

The following examinations must be completed, with content as described below for each item:

- Vital Signs
- Seizure Frequency and Type
- MoCA, RAVLT, QOLIE-31, BNT-60, BVMT-R, HAMD, BAI
- Adverse Events, Concomitant Medications/Treatments
- The Investigator may add the following examinations based on the subject's clinical presentation: Serum Pregnancy Test (WOCBP only), Physical Examination, Hematology, Blood Biochemistry, CRP, Coagulation Panel, Urinalysis, Stool Routine + Occult Blood, EEG, PRA, DSA, Cytokines, Immune Panel, Peripheral Blood T-cell Subsets, Tumor Markers

#### 8.2 Assessment Conment

##### 8.2.1 Safety Evaluation

1. AE/SAE

- Incidence and severity of AEs, SAEs, and treatment-related adverse events (TRAEs) associated with the implant and/or surgery during the treatment and follow-up periods.

2. Physical Examination

- Physical examination: Includes skin and mucous membranes, lymph nodes, head and neck, chest, abdomen, spine and extremities, musculoskeletal system, and neurological system.
- The Investigator should pay particular attention to clinical symptoms related to pre-existing severe conditions.

3. Vital Signs

- Vital signs will be measured after the subject has rested in a sitting or supine position for at least 5 minutes, including body temperature (axillary), heart rate (pulse rate), respiration, and blood pressure.

4. 12-Lead Electrocardiogram (ECG)

- A supine 12-lead ECG will be performed using an ECG device at time points specified in the Study Flowchart. The ECG device will automatically calculate heart rate and measure PR, QRS, QT, and QTc intervals.
- When ECG examinations overlap with vital sign measurements and biosample collection (e.g., peripheral venous blood, skin, etc.), the following priority order should be followed: 12-Lead ECG, vital signs, biosample collection.

5. Other Safety Assessments

- Laboratory tests (Hematology, Blood Biochemistry, C-Reactive Protein, Urinalysis, Stool Routine, Coagulation Panel, Electroencephalogram), Brain MRI, Immune Panel, Cytokines, Peripheral Blood T-cell Subsets, Tumor Screening (Tumor Markers, MRI, CT brain tumor imaging).

The Investigator must review laboratory reports. Laboratory reports must be filed with source documents. Clinically significant abnormal laboratory results only include laboratory abnormalities unrelated to the underlying disease, unless the Investigator judges that the subject's abnormal results are more severe than expected.

During participation in the study, all clinically significant abnormal laboratory results should undergo repeated measurements until the values return to normal, return to baseline values, or are deemed by the Investigator or medical expert to no longer be clinically significant.

1) If, within a period considered reasonable by the Investigator, values do not return to normal or baseline, the etiology needs to be investigated, and the medical expert must be notified.

2) All protocol-mandated laboratory assessments must be conducted according to the laboratory manual and study procedures.

3) If laboratory results from non-protocol-mandated tests conducted in the study site's local laboratory indicate a need for adjustments to the subject's management plan or are deemed clinically significant by the Investigator (e.g., SAE or AE or dose adjustment), then these laboratory results must be recorded in the eCRF.

##### 8.2.2 Efficacy Evaluation

1. Brain Tissue Metabolic and Functional State: Evaluate the change (improvement) in 18F-FDG uptake in the injection area compared to pre-transplantation baseline, assessed by FDG-PET at 6 and 12 months post-surgery.

2. GABA Levels: Evaluate the change (improvement) in GABA levels, related molecular levels and ratios, and structural changes in the injection area compared to pre-transplantation baseline, assessed by MRS at 1, 3, 6, 9, and 12 months post-surgery.

3. Responder Rate: Evaluate the change (improvement) in the percentage of subjects with a ⩾50% reduction in seizure frequency at 1-12 months post-surgery compared to pre-transplantation baseline.

4. Seizure Frequency: Evaluate the change (improvement) in subject seizure frequency (number of seizures per 28 days) at 1-12 months post-surgery compared to pre-transplantation baseline, based on seizure diaries and EEG; includes the frequency of all seizure events and disabling seizures.

5. Quality of Life Assessment: Evaluate the change (improvement) in QOLIE-31 scores at 1, 3, 6, 9, and 12 months post-surgery compared to pre-transplantation baseline.

6. Neurocognitive Function Assessment: Evaluate the change (improvement) in scores (or results) for MoCA, RAVLT, BNT-60, and BVMT-R at 1, 3, 6, 9, and 12 months post-surgery compared to pre-transplantation baseline.

7. Mood Disorder Assessment: Evaluate the change (improvement) in HAMD and BAI scores at 1, 3, 6, 9, and 12 months post-surgery compared to pre-transplantation baseline.

### 9 Management of Adverse Events

AEs will be reported by the subject (or, where appropriate, by a caregiver, proxy, or the subject's legally authorized representative) to the Investigator and any qualified designee. They will be responsible for detecting, documenting, and entering events that meet the definition of an AE or SAE.

#### 9.1 Definition of Adverse Event (AE)

An AE is defined as any untoward medical occurrence in a subject administered a study intervention (for this study, starting from the ALC05 cell injection surgery), but which does not necessarily have a causal relationship with the treatment. Thus, an AE can be any unfavorable and unintended sign (including an abnormal laboratory finding), symptom, or disease temporally associated with the use of the investigational product, regardless of its causal relationship with the product.

**Events that Meet the Definition of an AE:**

- Any abnormal laboratory test result (hematology, serum biochemistry, or urinalysis) or other safety assessment (e.g., ECG, radiological scan, vital sign measurement) that is clinically significant as judged by the Investigator (and not related to the progression of the underlying disease), including laboratory results that worsen from baseline;

- Worsening of a pre-existing chronic or intermittent condition, including an increase in the frequency and/or intensity of the condition;

- New medical conditions detected or diagnosed after the administration of the study treatment (even if the condition might have existed before the start of the study);

- Signs, symptoms, or clinical sequelae that may suggest a drug-drug interaction;

- Signs, symptoms, or clinical sequelae suggesting a suspected overdose while on study treatment or concomitant medication. Overdose itself will not be reported as an AE/SAE unless it was taken with suicidal/self-harm intent. For the latter, the event should be reported regardless of sequelae.

**Events Not Meeting the Definition of an AE:**

- Any clinically important abnormal laboratory test results or other abnormal safety assessments related to the underlying disease, unless the Investigator judges the subject's condition to be more severe than expected;

- The condition being treated by this study, or the expected progression, signs, or symptoms of that condition, unless the subject's condition is more severe than expected;

- Medical procedures or surgical interventions resulting from an AE (e.g., endoscopy, appendectomy);

- Situations where no untoward medical event occurred (social reasons and/or hospitalization for convenience);
- Daily fluctuations of pre-existing or current diseases/conditions that are present or detected at the start of the study without worsening.

##### 9.2 Definition of Serious Adverse Event (SAE)

If an event does not meet the above definition of an AE, then even if it meets the seriousness criteria, it cannot be classified as an SAE (e.g., hospitalization for symptoms of the disease under study, death due to disease progression).

A Serious Adverse Event is defined as any untoward medical occurrence that meets any of the following criteria:

a. **Results in Death**

b. **Is Life-Threatening**

The term "life-threatening" in the definition of "serious" refers to an event in which the subject was at risk of death at the time of the event, and not to an event which hypothetically might have caused death if it had occurred in a more severe form.

c. **Requires Inpatient Hospitalization or Prolongation of Existing Hospitalization**

Generally, inpatient hospitalization means that the subject is kept in a hospital or emergency ward (typically at least overnight) for observation and/or treatment because they cannot receive adequate observation and/or treatment on an outpatient basis or in a physician's office. Complications occurring during hospitalization are AEs. If a complication prolongs hospitalization or meets any other seriousness criterion, it is judged as "serious." When it is uncertain whether "hospitalization" occurred or was medically necessary, the AE should be considered "serious."

Elective hospitalization for a pre-existing condition that has not worsened from baseline will not be considered an AE.

d. **Results in Persistent or Significant Disability/Incapacity**

- Disability refers to a substantial disruption of a person's ability to conduct normal life functions;
- This definition does not include medically minor events such as uncomplicated headaches, nausea, vomiting, diarrhea, influenza, and accidental trauma (e.g., a sprained ankle), which may interfere with or hinder daily life but do not constitute a substantial disruption.

e. **Is a Congenital Anomaly/Birth Defect**

##### 9.2.1 Hospitalization/Prolongation of Hospitalization

AEs in clinical studies that lead to inpatient hospitalization (even if less than 24 hours) or prolongation of existing hospitalization should be considered SAEs. Hospitalization does not include the following:

- Rehabilitation facilities;
- Nursing homes;
- Routine emergency room admissions (less than 24 hours);
- Day surgery (e.g., outpatient/same-day/non-inpatient surgery);
- Admissions required for follow-up examinations.

#### 9.3 Transplant- and/or Procedure-Related Adverse Events (TRAEs)

TRAEs are AEs (serious or non-serious) concerning ALC05 or the study's scientific and medical focus that require continuous monitoring by the Investigator and immediate notification to the medical expert. In this study, TRAEs include: CRS (Cytokine Release Syndrome), infection, fever, allergic reactions, GvHD (Graft-versus-Host Disease), IEDs (Interictal Epileptiform Discharges), and primary malignancy.

#### 9.4 AE/SAE Severity

The severity of AEs in this study is graded according to the Common Terminology Criteria for Adverse Events (CTCAE v5.0). The grading of AE severity is as follows:

Grade 1: Mild; asymptomatic or mild; clinical or diagnostic observations only; intervention not indicated.

Grade 2: Moderate; minimal, local or noninvasive intervention indicated; limiting instrumental ADL (instrumental Activities of Daily Living include cooking, shopping for groceries or clothes, using the telephone, managing money, etc.).

Grade 3: Severe or medically significant but not immediately life-threatening; hospitalization or prolongation of hospitalization indicated; disabling; limiting self-care ADL (self-care Activities of Daily Living include bathing, dressing, eating, toileting, taking medications, and not being bedridden).

Grade 4: Life-threatening consequences; urgent intervention indicated.

Grade 5: Death related to AE.

#### 9.5 Causality Assessment of AE/SAE and Cell Therapy

Causality assessment is one of the criteria used in determining regulatory reporting requirements. The Investigator is responsible for assessing whether a reasonable possibility of a causal relationship exists between the study treatment and each AE/SAE that occurs.

"Reasonable possibility" of a causal relationship means that there are facts, evidence, and/or arguments suggesting a causal relationship, rather than merely the inability to rule out a causal relationship.

The Investigator will determine causality through clinical judgment, referencing the Investigator's Brochure (IB) and/or product information for marketed products. Other factors will be fully considered when assessing causality, such as underlying diseases, concomitant treatments, and other risk factors, as well as other events causally related to the investigational product.

For each AE/SAE, the Investigator must document in the clinical record that he/she has reviewed the AE/SAE and made a causality assessment. In some cases, the Investigator may not be able to provide sufficient other information regarding an SAE in the initial report to the medical expert. However, the Investigator is required to perform a causality assessment for each event before the initial report is submitted to the medical expert. The Investigator may update his/her causality assessment based on subsequent follow-up information but is required to submit an SAE follow-up report for this updated causality assessment.

The following definitions will be used for assessing the causal relationship between AEs and the investigational product in this study:

| 5-Grade Classification | Judgment Criteria |
|---|---|
| **Definitely Related** | The reaction type is consistent with the known reaction profile of the suspected drug. A reasonable temporal relationship exists after drug administration. The adverse reaction subsides or disappears upon dose reduction or discontinuation, and reappears upon re-administration. |
| **Probably Related** | The reaction type is consistent with the known reaction profile of the suspected drug. A reasonable temporal relationship exists after drug administration. The adverse reaction subsides or disappears upon dose reduction or discontinuation. However, the subject's clinical state or other factors could also potentially explain the reaction. |
| **Possibly Related** | The reaction type is consistent with the known reaction profile of the suspected drug. A reasonable temporal relationship exists after drug administration. The adverse reaction subsides or is not apparent upon |

| | |
|---|---|
| | dose reduction or discontinuation. However, the subject's clinical state or other factors could also explain the reaction. |
| **Possibly Unrelated** | The reaction type is not highly consistent with the known reaction profile of the suspected drug. The temporal relationship after drug administration is not highly reasonable. The subject's clinical state or other factors could also potentially explain the reaction. |
| **Unrelated** | The reaction type is inconsistent with the known reaction profile of the suspected drug. The temporal relationship after drug administration is not reasonable. The subject's clinical state or other factors can fully explain the reaction. The reaction subsides or disappears after ruling out clinical symptoms or other causes. |

##### 9.6 Follow-up of AE/SAE

After the initial AE/SAE report, the Investigator is required to proactively follow up on subject information during every subsequent subject visit/contact. All SAEs must be followed up until recovery or return to baseline or death or lost to follow-up.

- The Investigator has the obligation to perform or arrange for additional examinations and/or assessments, according to medical guidelines or clinical requirements, to clarify the nature and/or causality of the AE or SAE as fully as possible. For example, additional laboratory tests, histopathological examinations, or consultation with other healthcare professionals may be conducted.
- New or updated information must be recorded in the initially completed CRF.
- The Investigator will report updated information to the medical expert within 24 hours of receiving it.

##### 9.7 SAE Reporting Process

- In this study, data will be collected from the first dose until the end of follow-up.

  If a subject experiences an SAE during the trial, regardless of its relationship with the investigational medicinal product, the Investigator should immediately take appropriate therapeutic measures to ensure the subject's safety, and promptly report to the clinical trial responsible unit's head. Upon learning of an SAE, the Investigator must immediately notify the Sponsor and/or CRO relevant personnel. The medical personnel of the Sponsor and/or CRO will medically review the content of the SAE report form and provide feedback to the Investigator. After multi-party review and confirmation, the Investigator will sign and date the report.

For SAEs where information is temporarily incomplete or uncertain, the Sponsor and/or CRO relevant personnel should also be promptly notified. More information will be supplemented later in the form of a follow-up report. The narrative section of the SAE report should describe in detail:

a) SAE symptoms

b) Severity

c) Onset date and time

d) Resolution date and time

e) Measures taken

f) Follow-up date and time

g) Method of follow-up

h) Outcome

All SAEs should also be recorded in the eCRF. The information provided in the SAE report form must be consistent with the data recorded in the eCRF for that event.

After learning of an SAE, the Investigator will complete the SAE report, submit it for medical review according to the medically established SAE review process. Once the report is finalized, the Investigator/CRC will email the signed scanned copy to the relevant medical personnel within 24 hours. For reports involving death events, the Investigator must also provide other required documents, such as autopsy reports and the final medical report.

If the Investigator deems an SAE to be unrelated to ALC05 but potentially related to study conditions (e.g., discontinuation of previous treatment, or comorbidities during the trial), this relationship should be detailed in the narrative section of the SAE report form.

#### 9.8 Pregnancy

- Specific information on pregnancies of female subjects and, if necessary, female partners of male subjects will be collected from the start of the study until 12 months after the end of follow-up.

- If a pregnancy is reported, the Investigator must notify the medical expert of the pregnancy event within 24 hours.

- Abnormal pregnancy outcomes (e.g., spontaneous abortion, fetal death, stillbirth, congenital anomalies, and ectopic pregnancy) will be reported as SAEs.

### 10 Data Analysis and Statistical Considerations

#### 10.1 Sample Size Definition

This study plans to enroll 12 subjects. As this is an exploratory study with no formal statistical hypotheses, it is anticipated that enrolling 12 subjects will be sufficient to meet the study's objectives for exploring preliminary safety and efficacy.

#### 10.2 Statistical Analysis Methods

The statistical analysis sets include the Safety Analysis Set and the Full Analysis Set:

- Full Analysis Set (FAS):

  All enrolled subjects who received ALC05 treatment constitute the Full Analysis Set for this study. The FAS will be used for demographic and baseline characteristics analysis and efficacy analysis.

- Safety Analysis Set (SAS):

  All enrolled subjects with any safety evaluation data available constitute the Safety Analysis Set for this study. The SAS will be used for all safety data analyses.

- Per-Protocol Set (PPS):

  Subjects who strictly followed the clinical trial protocol and completed all treatments and follow-ups constitute the Per-Protocol Set for this study.

SAS® Version 9.4 or higher will be used for all statistical analyses. For quantitative data, mean, standard deviation, median, maximum, minimum, and the 95% confidence interval for the estimated mean will be presented. For categorical and ordinal data, frequency (proportion), rate, and the 95% confidence interval for the estimated rate will be presented.

Unless otherwise specified, continuous endpoint variables will be statistically described by the number of non-missing cases, mean, standard deviation, median, minimum, and maximum. Categorical endpoint variables will be described by counts and percentages. For time-to-event endpoints, descriptive analysis will include the number of non-missing cases, median time, quartiles, minimum, maximum, and the number of events/censored cases. For data collected at assessment time points, summaries for each time point and changes from baseline will be provided. Additionally, a complete listing of all analysis data will be provided by dose group, subject, and assessment time point, with outliers marked.

Unless otherwise specified, baseline in statistical analyses is defined as the last non-missing value prior to study treatment.

##### 10.2.1 Subject Distribution and Baseline Characteristics Analysis

For all enrolled subjects, the number and percentage of subjects entering and completing each study phase, as well as those who prematurely withdrew from each phase and the reasons for withdrawal, will be statistically described. The number of subjects included in each analysis set will be summarized by dose group.

Descriptive statistical analysis of demographic and baseline characteristics will be performed based on the Safety Analysis Set. A summary analysis of study drug exposure will be provided.

##### 10.2.2 Safety Analysis

Safety analysis will be performed based on the Safety Analysis Set. All safety endpoints will be summarized using descriptive statistics by dose group and for the total cohort.

TEAEs are defined as adverse events that occur or worsen from the time of study drug administration onward. AEs will be coded using the Medical Dictionary for Regulatory Activities (MedDRA) and graded according to CTCAE 5.0.

The number and incidence of various AEs will be summarized by System Organ Class (SOC), and the incidence of TRAEs as judged by the Investigator will be summarized by severity. The incidence rates of AE, SAE, and TRAE will be summarized.

For safety data collected at assessment time points, descriptive analysis of assessment data for each time point will be performed; if appropriate, a summary of changes from baseline will also be provided. For categorical assessment results, counts and percentages of subjects for each category, as well as cross-tabulations of severity grades or changes in clinical significance relative to baseline, will be provided.

##### 10.2.3 Efficacy Analysis

The primary analysis population for all efficacy endpoints is the FAS, which includes all randomized subjects and follows the ITT principle. Sensitivity analysis will be performed using the PPS, which consists of subjects meeting key protocol criteria without major deviations. All efficacy analyses will be performed by treatment group and for the overall study population. For continuous endpoints, summary statistics (n, mean, standard deviation, median, minimum, maximum) will be provided for each planned post-baseline visit/time point, and changes from baseline for each visit will be calculated and summarized. For categorical endpoints, counts and percentages will be listed by treatment group.

Descriptive summaries of response rates will include the number (n), percentage (%), and 95% Confidence Interval (CI) of responders, non-responders, and seizure-free subjects for each time period (M1-M3, M4-M6, M7-M9, M10-M12) and overall (M1-M12), summarized by treatment group. The 95% CI for percentages will be calculated using the Clopper-Pearson exact method. Inter-group comparisons will use Fisher's Exact Test (two-sided, $\alpha=0.05$) to test for differences in response rates (responder vs. non-responder) between treatment groups (Low-Dose vs. High-Dose). Odds Ratios (OR) and their associated 95% CIs will be reported.

Descriptive statistics for seizure frequency: For each time point and treatment group, summary statistics will be calculated for absolute seizure frequency, change from baseline, and percentage change from baseline. Summary statistics will include: number of subjects (n), Mean ± Standard Deviation (SD), Median, First Quartile (Q1), Third Quartile (Q3), Minimum (Min), Maximum (Max), and the 95% Confidence Interval (CI) of the mean. Intra-group analysis will test whether changes in seizure frequency from baseline at each time point (M1 to M12) are statistically significant. A paired t-test will be used for normally distributed data; the Wilcoxon signed-rank test will be used for non-normally distributed data. P-values and 95% CIs for mean change will be reported. Inter-group comparison will analyze the difference in absolute change from baseline between the Low-Dose and High-Dose groups at each time point. An independent samples t-test will be used for normally distributed data, and the Wilcoxon rank-sum test will be used for non-normally distributed data. The mean difference, its 95% CI, and descriptive p-values will be reported.

Descriptive statistics for scale analysis will include subscale scores and total scores. Scores at baseline and all post-baseline visits will be summarized by treatment group. Changes in scores from baseline will be calculated and summarized at each time point. Summary statistics will include: n, mean ± SD, median, Q1, Q3, Min, Max, and 95% CI. Responder analysis will define clinically meaningful improvement as an increase in total score from baseline of ⩾X points (defined according to the Minimal Clinically Important Difference [MCID] for different scales). The number (n) and percentage (%) of subjects achieving this clinically meaningful improvement will be calculated, and associated 95% CIs determined using the Clopper-Pearson exact method. Intra-group analysis will use paired t-tests at each post-baseline visit to test for statistical differences in subscale and total score changes from baseline. Effect size (Cohen's d) will be calculated to assess the magnitude of efficacy. Inter-group comparison will use Analysis of Covariance (ANCOVA) models at each visit to compare the "change in score from baseline" between treatment groups: Change from Baseline = Treatment Group + Baseline Score. The Least Squares (LS) mean difference between groups, its 95% CI, and descriptive p-values will be reported. Specific statistical analysis methods are detailed in the Statistical Analysis Plan (SAP).

##### 10.2.4 Data and Safety Monitoring

This study will establish a DSMB, responsible for evaluating the safety of the intervention during the trial and overseeing the overall implementation of the clinical trial. The DSMB will provide recommendations regarding the termination or continuation of the clinical trial. To help enhance the integrity of the trial, the DSMB may provide recommendations on subject screening, recruitment and retention, compliance management, and data management and quality control processes.

The DSMB's scope of responsibilities also covers critical operational decisions, including reviewing unblinded safety data after the first subject in each of the low and high dose groups completes at least 3 months of safety observation. Based on this review, the DSMB will formally assess whether the safety results meet expectations, thereby deciding whether to approve the simultaneous entry and randomization of the subsequent 10 subjects into both dose groups.

Additionally, the DSMB is responsible for reviewing all protocol amendments related to safety, continuously and regularly reviewing aggregated safety data, assessing the overall benefit-risk balance of the study, and making recommendations regarding the continuation, protocol amendment, or termination of the study based on pre-defined stopping rules. In fulfilling its duties, particularly in handling unblinded data, the DSMB will strictly ensure that the study team remains blinded to treatment assignments at all times to maintain the objectivity and scientific validity of the trial. The board will also hold regularly scheduled meetings for reviewing and adjudicating all SAEs, and may convene ad hoc meetings if necessary.

The DSMB will consist of the study Investigator, the Sponsor-designated Medical Monitor, the Sponsor's Medical Monitor, the study statistician, and three independent physicians with clinical expertise in epilepsy and cell therapy who are not involved in the study's implementation and are blinded to treatment assignments. The committee will conduct routine reviews of the trial's implementation process and the accumulating safety and efficacy data. To facilitate independent unblinded data review, an independent statistician will be designated to handle the statistical tables and listings of the study data.

The three independent and blinded physicians will be the voting members of the committee, who will collectively decide whether to continue or stop further subject enrollment. The Sponsor's Medical Monitor, the study statistician, the independent statistician, and other committee members will be non-voting members.

The DSMB Charter will clearly define the committee's responsibilities and meeting procedures. This charter must be finalized before the enrollment of the first patient. The DSMB will operate according to the procedures outlined in the independently documented DSMB Charter.

### 11 Data Management

#### 11.1 Data Entry

Data entry for this study will be performed via direct entry into the EDC system. The Investigator will accurately, timely, completely, and consistently enter data into the EDC system according to the EDC filling guidelines formulated by the data management department, based on the subject's source document information.

#### 11.2 Database Verification and Query Management

Data verification is not limited to the data management CRO but should be led by the CRO's data management personnel, with collaborative efforts from clinical monitors, medical personnel, and others. Clinical monitors are responsible for verifying the consistency between source documents and data entered into the EDC, ensuring data authenticity and completeness. Data management personnel are responsible for performing data verification through the EDC system by configuring logical check rules, manual reviews, and external data consistency checks, ensuring data logic and accuracy. Medical personnel will perform medical review of certain data using their professional medical knowledge, ensuring data scientific validity. If necessary, statisticians and SAS programmers will provide professional technical support for data verification operations, to improve verification efficiency, such as identifying outliers in the data.

Data issues found by each verification module during data verification will be addressed by the personnel responsible for that module, who will create data queries within the EDC system. These queries will be answered by the Investigator. If the Investigator's response resolves the query, the verifier will close the query; if the query is not resolved, new queries may be created until the query is resolved or reasonably explained.

##### 11.3 Database Lock and Submission

According to the database lock procedure, once all pre-lock steps are completed, the database lock should be formally approved in writing, and data editing permissions for the database should be revoked, leading to database lock. If issues are found after database lock that require modification, and if it is necessary to unlock the database for modification, the unlock and re-lock procedures must be strictly followed.

#### 12 Risks and Mitigation

Existing Clinical Research Results:

ALC05 cell product is an investigational product, and clinical safety data for this product are not yet available.

Based on similar clinical studies, the potential safety risks during the ALC05 transplantation phase are as follows.

##### 12.1 Product Tumorigenicity Risks and Mitigation

Stem cells share some biological characteristics with tumor cells, such as unlimited proliferation potential, insensitivity to apoptotic signals, and similar growth regulatory mechanisms. Preclinical animal model studies indicate that undifferentiated human embryonic stem cells or iPSCs carry a risk of forming benign or even malignant teratomas after transplantation.

However, ALC05 used in this study does not consist of undifferentiated stem cells; rather, it is comprised of high-purity GABAergic interneurons obtained through directed differentiation. Quality control testing (flow cytometry) of the final product showed no detectable iPSC-specific biomarkers, indicating that the residual level of undifferentiated iPSCs in the final product is extremely low. Theoretically, with the elimination of residual undifferentiated iPSCs, the tumorigenic risk of the product is significantly reduced.

Furthermore, no tumors were observed in prior animal studies: a 6-month tumorigenicity study of iPSC-derived GABAergic interneurons (ALC05) in immunodeficient mice revealed no abnormal structures within the hippocampal transplant area.

Management Measures: Regular imaging examinations such as MRI and CT scans, along with the monitoring of tumor markers such as AFP (alpha-fetoprotein) and CEA (carcinoembryonic antigen), can allow for the early detection of potential tumor risks.

##### 12.2 Graft-Versus-Host Disease (GvHD) Risks and Mitigation

ALC05 cells are low-immunogenicity iPSC-derived GABAergic interneurons. There is a risk of developing GvHD following ALC05 cell transplantation. If a GvHD event occurs, investigators can manage the subject by referring to the NCCN Guidelines for Patients®: Graft-Versus-Host Disease.[31]

##### 12.3 Stereotactic Injection Surgery Risks and Mitigation

Stereotactic injection surgery is a minimally invasive surgical method that combines image-guidance technology to accurately target areas within the brain for drug or cell injection. By planning the surgical trajectory using fused imaging such as MRI and CT, it ensures that the guide needle or minimally invasive instruments accurately reach the predetermined target. It is widely used in the treatment of neurological diseases such as epilepsy and Parkinson's disease. As a critical step in epilepsy cell therapy, despite its high precision and minimally invasive nature, this surgery carries potential risks including hemorrhage, nerve damage, infection, and misplacement, which could lead to neurological dysfunction or other serious adverse reactions.

To address these risks, we have implemented several mitigation measures: First, high-precision image guidance and navigation technologies are employed to ensure the accuracy of the needle trajectory; second, the procedure is performed by an experienced professional team strictly adhering to aseptic principles to reduce the occurrence of hemorrhage and infection. Additionally, real-time monitoring of neural responses and imaging confirmation during surgery help avoid damaging adjacent critical structures. Close observation of the patient's condition post-operatively ensures early detection and management of potential complications, thereby safeguarding surgical safety and treatment efficacy.

##### 12.4 Immunosuppression Side Effects and Mitigation

In this study, subjects will receive immunosuppressive therapy to prevent or reduce immune rejection following ALC05 cell transplantation. The selected immunosuppressant is tacrolimus, whose potent immunosuppressive effect is crucial for maintaining cell survival. However, like all potent immunosuppressants, the use of tacrolimus is associated with a range of potential adverse reactions. The following outlines the common and serious adverse reactions of tacrolimus and details the risk mitigation strategies of this study.

Adverse reactions to tacrolimus are generally dose-related and mostly reversible, subsiding or disappearing upon dose reduction. Due to the complexity of the patients' underlying diseases and concomitant medications, it is difficult to definitively establish a direct correlation between all adverse reactions and the immunosuppressant. The primary categories of adverse reactions are as follows:

- Infections and Infestations (Very Common): As with other potent immunosuppressants, patients taking tacrolimus have a significantly increased risk of viral (e.g., CMV, BK virus-associated nephropathy, JC virus-associated progressive multifocal leukoencephalopathy [PML]), bacterial, fungal, and protozoal infections. Pre-existing infections may worsen, and systemic or localized infections may occur.

- Benign, Malignant, and Unspecified Neoplasms (incl cysts and polyps) (Common): Patients receiving immunosuppressive therapy have an increased risk of developing malignancies. Benign and malignant tumors associated with tacrolimus treatment have been reported, including Epstein-Barr virus (EBV)-associated lymphoproliferative disorders, skin malignancies, and Kaposi's sarcoma.

- Metabolism and Nutrition Disorders (Very Common): Hyperglycemic conditions (including diabetes) and hyperkalemia are very common. Also common are hypomagnesemia, hypophosphatemia, hypokalemia, hypocalcemia, hyponatremia, fluid overload, hyperuricemia, decreased appetite, anorexia, metabolic acidosis, hyperlipidemia, hypercholesterolemia, hypertriglyceridemia, and other electrolyte abnormalities.

- Nervous System Disorders (Very Common): Tremor and headache are very common. Common neurological adverse reactions include seizures, disturbances in consciousness, paresthesia and dysesthesia, peripheral neuropathies, dizziness, dysgraphia, and neurological disorders. Rare occurrences include coma, central nervous system hemorrhage, cerebrovascular accidents, paralysis, encephalopathy, and amnesia.

- Cardiovascular Disorders: Hypertension is very common. Ischemic coronary artery disorders, tachycardia, hemorrhage, thromboembolic and ischemic events, peripheral vascular disorders, and vascular hypotensive disorders are also common.

- Hepatobiliary Disorders (Common): Cholestasis and jaundice, hepatocellular damage and hepatitis, and cholangitis are common. Hepatic artery thrombosis, veno-occlusive liver disease, and extremely rarely, hepatic failure can occur.

- Renal and Urinary Disorders (Very Common): Renal impairment is very common. Renal failure, acute renal failure, oliguria, renal tubular necrosis, toxic nephropathy, and urinary system disorders are common.

- Blood and Lymphatic System Disorders (Common): Anemia, leukopenia, thrombocytopenia, leukocytosis, and abnormal red blood cell analyses are common. Coagulation disorders and pancytopenia are rare.

- Psychiatric Disorders (Very Common): Insomnia is very common. Anxiety symptoms, confusion, disorientation, depression, depressed mood, mood disorders, nightmares, hallucinations, and mental disorders are common.

- Skin and Subcutaneous Tissue Disorders (Common): Pruritus, rash, alopecia, acne, and hyperhidrosis are common.

- Other Common Adverse Reactions: Asthenic conditions, febrile disorders, edema, pain, and discomfort.

To minimize the risks associated with tacrolimus and ensure subject safety, this study will implement the following strict risk control and management measures:

- Inclusion/Exclusion Criteria: Strict adherence to inclusion and exclusion criteria to exclude patients with a history of significant infection, malignancy, severe hepatic or renal insufficiency, or coagulation abnormalities, thereby reducing the probability of adverse reactions in high-risk subjects.

- Dosing and Titration: Cautious dose titration based on individual differences, Therapeutic Drug Monitoring (TDM) results, and clinical response to achieve the minimum effective and safe dose.

- TDM: Regular monitoring of tacrolimus trough concentrations to maintain levels within the target therapeutic window, optimizing efficacy while minimizing toxicity.

- Strict Infection Monitoring: Close monitoring of subjects for signs of infection (e.g., fever, cough, chills), with regular testing of inflammatory markers such as CBC and CRP. Upon any sign of infection, immediate etiological diagnosis and aggressive, appropriate anti-infective treatment will be initiated.

- Tumor Screening: Long-term immunosuppression may increase the risk of malignancy. Subjects will undergo regular tumor-related examinations and follow-ups during the study.

- Metabolic Parameter Monitoring: Regular monitoring of metabolic indicators including blood glucose, potassium, magnesium, phosphorus, calcium, sodium, uric acid, and lipids. Interventions (e.g., dietary guidance, medication adjustment) will be implemented promptly based on results.

- Hepatic and Renal Function Monitoring: Regular assessment of liver function (ALT, AST, ALP, GGT, TBIL, etc.) and renal function (serum creatinine, blood urea nitrogen, eGFR, etc.) to evaluate tacrolimus-induced hepatotoxicity and nephrotoxicity.

- Neuropsychiatric Symptom Monitoring: Close observation of the subject's neurological symptoms (e.g., tremor, headache) and psychiatric state (e.g., insomnia, anxiety, confusion), utilizing standardized scale assessments for early detection and intervention.

- Hematological Monitoring: Regular complete blood counts (CBC) to monitor hematological adverse reactions such as anemia, leukopenia, and thrombocytopenia.
- Drug Interactions: Detailed recording of all concomitant medications, maintaining vigilance against interactions between tacrolimus and CYP3A4 inhibitors/inducers, calcium channel blockers, etc., to avoid increased toxicity or reduced efficacy.
- Patient Education: Comprehensive medication education for subjects and their families, informing them of potential adverse reactions, the importance of monitoring, and when to seek medical assistance.
- Pain Management: Special adverse reactions such as Calcineurin-Inhibitor Induced Pain Syndrome (CIPS) will be closely monitored, and adjustment of the tacrolimus dose will be considered based on clinical presentation.

##### 12.5 Donor Safety Risks and Mitigation

ALC05, used in this study, is a cell suspension product prepared in vitro from the blood of healthy adult donors. To mitigate potential risks originating from the donor, we have conducted strict donor eligibility determinations for all donors through detailed examinations and tests. The examinations include a thorough understanding of the donor's medical and family history, as well as comprehensive laboratory tests. The laboratory tests encompass screening for potential infectious diseases (HIV, HBV, HCV, HTLV, and Syphilis). Only healthy adults who test negative for all parameters are eligible to donate. In addition to routine laboratory tests, donors undergo karyotype analysis and whole-genome sequencing to rule out the possibility of hereditary tumors.

The testing items refer to the third edition of FDA 21 CFR 1271 regulations.
